# Predicting One-Year CPAP Adherence and Use Trajectories Utilizing Baseline and Early Use Data

**DOI:** 10.64898/2026.10.01.26363483

**Authors:** Nathanael H. Hwang, Soniya Mishra, M. Brandon Westover, Robert J. Thomas, Emmanuel Mignot, Umakanth Katwa, Jiaxiao Shi, Rui Yan, Kendra A. Becker, Dennis Hwang, Matthew T. Klimper, Jessica Jara, Damien R. Stevens, Matthew K.P. Gratton, Diego R. Mazzotti, Haoqi Sun

**Author notes:** Diego R. Mazzotti and Haoqi Sun are co-senior authors. **Corresponding Author:** Nathanael Hwang, Beth Israel Deaconess Medical Center DA-0815, East Campus 330 Brookline Ave, Boston, MA 02215.

## Abstract

**Introduction:** Maintaining long-term positive airway pressure (CPAP) adherence is a major challenge when treating obstructive sleep apnea (OSA). We evaluated factors associated with one-year CPAP adherence and developed models predicting Month-12 adherence and CPAP-use trajectories.

**Methods:** CPAP-naive adults with OSA (apnea-hypopnea index [AHI]≥5) at Kaiser Permanente Southern California with 364-day observation period were analyzed (development/internal-validation cohort, n=14,906; temporal holdout, n=1,676). Logistic regression evaluated baseline associations with Month-12 adherence (mean≥4 hours/night during days 331-360). XGBoost models utilized *baseline* features (clinical, sleep-study, questionnaire data), *early CPAP-only* data (days 1-7), or *combined* to predict long-term CPAP use. A latent-class model identified trajectories over days 8-364. Predictive models were evaluated in both random and temporal holdouts.

**Results:** About one-third (34.6%) of the primary cohort (49.7±13.8 years, 63.2% male) were adherent during Month-12. Higher AHI and longer self-reported sleep duration favored adherence while greater comorbidity burden, depression, and substance-use disorder were associated with lower adherence. In held-out testing, adherence prediction using *baseline* features yielded an AUROC of 0.678. Using *CPAP-only* data, AUROCs were higher from 0.787 with seven days of usage data to 0.816 and 0.854 with 14 and 30 days. Models combining CPAP and baseline features increased Day-7 AUROC to 0.810. We identified four one-year trajectories: Early-Decliners (42.9%), Gradual-Decliners (17.4%), Stable-Moderate (19.0%), Stable-High (20.7%). Models predicting patient trajectory revealed macro-average AUROCs of 0.619, 0.756, and 0.770 for baseline, CPAP-only, and combined models.

**Conclusion:** Baseline pAbstractTextredictors may identify strategies for adherence support, while early CPAP data substantially improved predictive performance. These models may guide personalized follow-up but should not restrict CPAP access. External and implementation validation are needed.

## Introduction

Continuous positive airway pressure (CPAP) is an efficacious treatment for obstructive sleep apnea (OSA), but its clinical effectiveness depends on sustained use.^1^ Greater CPAP use is associated with improvements in sleepiness, functional status, and other OSA-related outcomes.^2^ Nevertheless, many patients use CPAP inconsistently or discontinue treatment over time.^3^ Although adherence is commonly assessed during the first 90 days of therapy, early adherence does not fully characterize whether treatment will be sustained over the longer term.^4,5^

Understanding factors associated with long-term CPAP adherence may identify opportunities to improve treatment delivery and to personalize adherence support. Demographic, socioeconomic, clinical, behavioral, and OSA-related characteristics have all been associated with variations in CPAP adherence, including age, OSA severity, sleepiness, comorbidity, and psychosocial factors.^6–12^ However, findings have varied across studies, and the extent to which pretreatment information can meaningfully predict long-term adherence remains uncertain. Identifying factors independently associated with adherence may also reveal sub-populations or modifiable factors that could potentially support more personalized treatment decisions and allocation of support resources before CPAP is initiated.

After treatment initiation, actual CPAP-usage data provides an additional source of information. Early use during the first days to weeks of therapy is strongly associated with subsequent adherence and may be more predictive than pretreatment characteristics.^13–19^ However, it remains uncertain how baseline clinical information and early treatment behavior interact. Furthermore, adherence is a longitudinal behavior that may evolve through distinct patterns of sustained use, gradual decline, or early discontinuation, patterns that may not be captured by a single adherence threshold.^20–26^

We therefore examined long-term CPAP adherence in a large, diverse clinical cohort using linked electronic health records (EHR), sleep study, questionnaire, and wirelessly-acquired CPAP data. We organized the study into four phases: (1) an epidemiologic evaluation of baseline factors associated with adherence; (2) prediction of Month-12 adherence outcomes using baseline (pre-treatment) information, early CPAP use, and both; (3) identification and prediction of one-year CPAP-use trajectories; and (4) development of reduced-feature model specifications for rapid implementation in new settings with less data availability.

## METHODS

### Study design and reporting

We conducted a retrospective cohort study within Kaiser Permanente Southern California (KPSC), an integrated health care system. KPSC comprises a network of 14 medical centers with collaborative but independent sleep medicine practices. CPAP devices are prescribed as a purchase and access to equipment is not dependent nor retrieved based on adherence status. The standard post-CPAP care during the period of the study was passive in which follow-up encounters were initiated by patients and not proactively performed with structured protocols. The study linked EHR, diagnostic sleep study, sleep-questionnaire, and CPAP data. Reporting was structured to follow the TRIPOD framework for multivariable prediction models and the STROBE framework for observational studies.^27,28^ Eligible index sleep studies occurred between February 22, 2016, and February 27, 2025, and CPAP setup dates occurred between February 22, 2016, and March 3, 2025. The study was conducted as part of the Human Sleep Project (HSP) under protocols approved by the BIDMC Institutional Review Board (#2024P000804/#2022P000417), which granted a waiver of informed consent for this retrospective study.

### Participants and analytic cohorts

Adults aged 18 years or older were eligible if they had: obstructive sleep apnea, defined by an apnea-hypopnea index scored with a 4% oxygen-desaturation criterion (AHI4) of at least 5 events/hour; prescribed CPAP therapy (AutoCPAP; Airsense 10/11, Resmed Corp); had at least one recorded CPAP-use day (to confirm active wireless transmission); and had a dated index sleep study and a complete 364-day CPAP observation window. We excluded patients: treated with bilevel PAP; with observed CPAP use before the recorded setup date (to eliminate those not CPAP-naïve); and with study-to-setup latency longer than 1,095 days (chart review indicated that patients up to that latency were still almost all primary initiations and not device replacements). If a patient had multiple sleep studies, the earliest sleep study date (and corresponding questionnaire if available) was selected as the sleep study index date.

Two baseline cohorts were used: (1) the *Questionnaire-Complete* cohort comprised 14,906 earlier-date patients and 1,676 patients in a temporal holdout. Eligibility for this cohort required completion of an extensive sleep intake questionnaire **(Supplement S1)**; (2) the *EHR-Sleep-Study* cohort comprised 74,462 earlier-date patients and 5,013 patients in a temporal holdout. All Questionnaire-Complete patients were nested within the EHR-Sleep-Study cohort which additionally included patients without a complete set of questionnaire data (any partially available questionnaire data was ignored). The Questionnaire-Complete cohort was the primary KPSC development and internal-validation cohort for all four phases unless use of the EHR-Sleep-Study cohort is specified.

The temporal holdout consisted of patients whose index sleep-study dates fell within the most recent six months of the observed study period (August 27, 2024, or later). For each binary outcome, the primary cohort was randomly divided into stratified 80% training and 20% testing samples.

### PAP measures and outcomes

Nightly CPAP records were aligned to days 1 to 360 for most analyses (and up to day 364 for Phase 3 trajectory analysis). Nights without recorded use were imputed zero minutes in outcomes. Centers for Medicare & Medicaid Services (CMS) adherence indicator was defined as use for at least 4 hours/night on at least 70% of nights during a consecutive 30-day interval within the first 90 days after setup date.

The prespecified *primary outcome* was mean CPAP use ≥4 versus <4 hours/night during days 331–360 after setup (the Month-12 window), calculated across all 30 calendar nights. *Secondary outcomes* were: (1) mean Month-12 use ≥2 hours/night; (2) any recorded use during Month 12 (with no use during all 30 nights labelled as *treatment abandonment*); and (3) CMS adherence.

### Phase 1: Epidemiologic evaluation of baseline factors and CPAP adherence

Phase 1 evaluated associations between pre-CPAP baseline features and adherence. Prespecified features included age, sex, race and ethnicity, geocoded area-level socioeconomic measures, smoking, anthropometrics, comorbidities, health care utilization and engagement, medication-related measures, diagnostic sleep-study features, and patient-reported sleep and symptom measures. Diagnoses recorded before the index sleep study were mapped to non-overlapping Charlson and Elixhauser components. Complete feature definitions are provided in **Supplement S2**.

The primary epidemiologic analysis used a prespecified 32-term multivariable logistic-regression model in the Questionnaire-Complete earlier-date cohort (n=14,906), with Month-12 mean use ≥4 versus <4 hours/night as the dependent variable. The model jointly included AHI, oximetry, diagnostic study-to-CPAP-setup latency, demographic and socioeconomic covariates, smoking, Charlson comorbidity burden, depression, substance use, body mass index, health care engagement, medication measures, and selected questionnaire items. Continuous effects are reported per 1-SD increment. Wald 95% confidence intervals were calculated, with Benjamini-Hochberg adjustment across prespecified exposure terms. Adjusted odds ratios were calculated for these features.

### Phase 2: Prediction of Month-12 and secondary adherence outcomes

Phase 2 evaluated prediction of the primary Month-12 outcome (≥4 versus <4 hours/night) and the three secondary outcomes in the Questionnaire-Complete cohort. We compared XGBoost models: (1) *baseline* (pre-treatment) models utilizing EHR, sleep-study, and questionnaire variables; (2) *CPAP-data only* (post-treatment) models utilizing seven CPAP-data elements after each daily landmark through 7 (i.e., day 1, days 1-2, days 1-3, etc.) and over days 1-14 and 1-30 beginning with the first night of actual CPAP use; and (3) *combined* baseline + CPAP data at each daily landmark through day 7. The CPAP-data features were mean hours per calendar night, percentage of nights used, percentage of nights with ≥4 h of use, mean duration on used nights, mean 95th-percentile leak, mean 95th-percentile pressure, and mean residual AHI. Setup-date-anchored windows were examined as a sensitivity analysis **(Supplement S3)**.

The baseline-only model used 156 raw variables, CPAP-only models used 7 variables, and combined models used 163 raw variables. XGBoost handled missing values natively. Hyperparameters were selected within the training sample using five outer and five inner stratified folds and 40 Optuna trials per outer fold^29^, and the median outer-fold parameters were used to refit the full training sample. Baseline logistic regression used standardized continuous variables, prespecified categorical reference levels, removal of zero-variance and collinear terms, variance-inflation-factor pruning at 10, missingness indicators for informatively missing medication variables, and distance-weighted five-nearest-neighbor imputation for other incomplete continuous predictors.

Discrimination was summarized by AUROC in pooled out-of-fold predictions, the random held-out test sample, and the temporal holdout, with 2,000-resample stratified bootstrap 95% confidence intervals. SHAP values in the held-out test sample described global feature attribution for the baseline-only, CPAP-data, and combined data for the primary outcome.^30,31^ Decision-curve analysis compared baseline XGBoost and logistic-regression models for Month-12 nonadherence (<4 hours/night).^32^ Temporal calibration was assessed for the three models.

### Phase 3: Trajectory identification and prediction

Phase 3 used a common day-8 landmark in the Questionnaire-Complete cohort. Calendar days 1-7 were defined from the first night with positive recorded CPAP use and reserved for prediction. The trajectory target was weekly mean hours per calendar night over days 8-364 (51 complete, nonoverlapping 7-day blocks). This nonoverlapping design ensured that no CPAP measurements (during days 1-7) used as predictors would contribute to the target trajectories.

A cubic Gaussian latent-class trajectory model was selected from one-through eight-class candidates in a 5,000-patient random sample training-only screen using convergence, Bayesian information criterion, minimum class size, posterior-assignment diagnostics, trajectory shape, and clinical distinctness.^33^ The selected four-class model was refitted in all 11,925 training patients. The frozen parameters from the latent model were applied to assign the remaining 2,981 held-out test and 1,676 temporal-holdout patients. Entropy, average posterior probability of assignment, and odds of correct classification assessed assignment quality.

Three multiclass XGBoost models were assessed to predict the four-class assignment: baseline-only, CPAP-data only, and combined. All models used the same nested five-outer-fold by five-inner-fold protocol with 40 tuning trials per outer fold and a final training-only fit. We report macro and class-specific one-versus-rest AUROCs for Early-Decliners, Stable-Moderate use, Gradual-Decliners, and Stable-High users, with 2,000-resample stratified bootstrap 95% confidence intervals. SHAP values in the random held-out test sample summarized multiclass feature attribution for the baseline-only and combined models. For feature domain attribution, native XGBoost contributions were averaged in absolute value across the four class margins, and feature-level mean absolute values were summed within sleep-study, EHR, questionnaire, and CPAP-data domains and divided by the sum across all encoded features. Temporal calibration was assessed one versus rest for each trajectory class in all three models. Conditional pairwise discrimination was assessed for all six trajectory-class pairs in the random held-out test and temporal holdout samples by restricting evaluation to patients in each pair and renormalizing the corresponding predicted probabilities.

### Phase 4: Reduced-Feature Models and Implementation Considerations

Phase 4 evaluated reduced-feature model specifications intended to support implementation in settings with different data availability in anticipation of a planned external validation. In addition to the full baseline, CPAP-data-only, and combined models from Phases 2 and 3, we developed two reduced-feature baseline families. The *parsimonious model* used 13 raw variables (21 encoded columns), selected based on data availability, importance in the KPSC models, and clinician judgment. The *Questionnaire-Sleep-Study* model used 98 raw variables: 76 questionnaire variables, 18 sleep-study variables, and age, sex, height, and weight; no other EHR-derived features were included. The same seven-feature CPAP-data-only model served as the common comparator. Complete reduced-feature definitions are provided in **Supplement S4**. The reduced-feature models predicted the same four Phase 2 outcomes and the frozen Phase 3 trajectory assignments. We used the same preprocessing, nested XGBoost tuning, KPSC training and held-out testing, and 2,000-resample bootstrap procedures described above.

### Statistical analysis

Continuous characteristics are summarized as mean (SD) when approximately symmetric and median [interquartile range] when skewed; categorical variables are summarized as number (percentage). Discrimination was quantified by the area under the receiver-operating-characteristic curve (AUROC) in pooled out-of-fold predictions, the random test sample, and the temporal holdout. Percentile 95% confidence intervals used 2,000 stratified bootstrap resamples. Hypothesis tests were two-sided with α=.05. Calibration-in-the-large and calibration slope used predicted log-odds as an offset and covariate, respectively. Brier scores used unrecalibrated probabilities; calibration curves used 10 equal-frequency groups.

## RESULTS

### Cohort characteristics and outcomes

Baseline characteristics for the Questionnaire-Complete and EHR-Sleep-Study cohorts are shown in Table 1. Among the 14,906 Questionnaire-Complete patients, mean age was 49.7±13.8 years, 9,419 (63.2%) were male, mean body mass index (BMI) was 35.8±8.4 kg/m², and mean AHI was 34.1±26.0 events/hour. Race and ethnicity distribution was 36.9% Hispanic, 6.1% Black, 12.6% Asian, and 37.5% White. Median study-to-setup latency was 15 [interquartile range, 9–28] days. The Questionnaire-Complete cohort was nested within the EHR-Sleep-Study cohort, and both demonstrated similar characteristics.

**Table 1.**
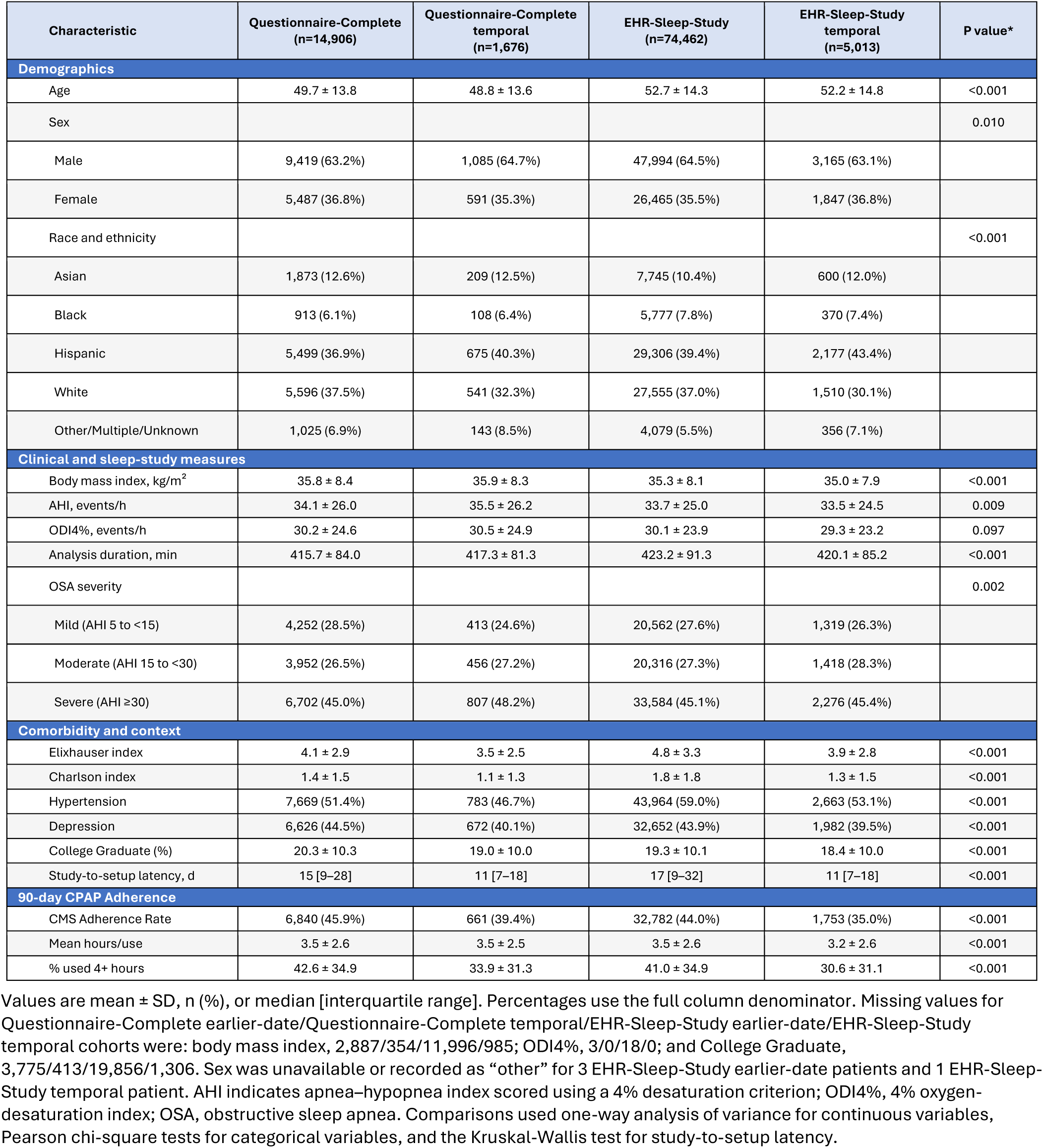
Baseline characteristics of the Questionnaire-Complete and EHR-Sleep-Study cohorts. Values are mean ± SD, n (%), or median [interquartile range]. Percentages use the full column denominator. Missing values for Questionnaire-Complete earlier-date/Questionnaire-Complete temporal/EHR-Sleep-Study earlier-date/EHR-Sleep-Study temporal cohorts were: body mass index, 2,887/354/11,996/985; ODI4%, 3/0/18/0; and College Graduate, 3,775/413/19,856/1,306. Sex was unavailable or recorded as “other” for 3 EHR-Sleep-Study earlier-date patients and 1 EHR-Sleep-Study temporal patient. AHI indicates apnea–hypopnea index scored using a 4% desaturation criterion; ODI4%, 4% oxygen-desaturation index; OSA, obstructive sleep apnea. Comparisons used one-way analysis of variance for continuous variables, Pearson chi-square tests for categorical variables, and the Kruskal-Wallis test for study-to-setup latency.

| Characteristic | Questionnaire-Complete<br>(n=14,906) | Questionnaire-Complete<br>temporal<br>(n=1,676) | EHR-Sleep-Study<br>(n=74,462) | EHR-Sleep-Study<br>temporal<br>(n=5,013) | P value* |
| --- | --- | --- | --- | --- | --- |
| **Demographics** |  |  |  |  |  |
| Age | 49.7 ± 13.8 | 48.8 ± 13.6 | 52.7 ± 14.3 | 52.2 ± 14.8 | <0.001 |
| Sex |  |  |  |  | 0.010 |
| Male | 9,419 (63.2%) | 1,085 (64.7%) | 47,994 (64.5%) | 3,165 (63.1%) |  |
| Female | 5,487 (36.8%) | 591 (35.3%) | 26,465 (35.5%) | 1,847 (36.8%) |  |
| Race and ethnicity |  |  |  |  | <0.001 |
| Asian | 1,873 (12.6%) | 209 (12.5%) | 7,745 (10.4%) | 600 (12.0%) |  |
| Black | 913 (6.1%) | 108 (6.4%) | 5,777 (7.8%) | 370 (7.4%) |  |
| Hispanic | 5,499 (36.9%) | 675 (40.3%) | 29,306 (39.4%) | 2,177 (43.4%) |  |
| White | 5,596 (37.5%) | 541 (32.3%) | 27,555 (37.0%) | 1,510 (30.1%) |  |
| Other/Multiple/Unknown | 1,025 (6.9%) | 143 (8.5%) | 4,079 (5.5%) | 356 (7.1%) |  |
| **Clinical and sleep-study measures** |  |  |  |  |  |
| Body mass index, kg/m<sup>2</sup> | 35.8 ± 8.4 | 35.9 ± 8.3 | 35.3 ± 8.1 | 35.0 ± 7.9 | <0.001 |
| AHI, events/h | 34.1 ± 26.0 | 35.5 ± 26.2 | 33.7 ± 25.0 | 33.5 ± 24.5 | 0.009 |
| ODI4%, events/h | 30.2 ± 24.6 | 30.5 ± 24.9 | 30.1 ± 23.9 | 29.3 ± 23.2 | 0.097 |
| Analysis duration, min | 415.7 ± 84.0 | 417.3 ± 81.3 | 423.2 ± 91.3 | 420.1 ± 85.2 | <0.001 |
| OSA severity |  |  |  |  | 0.002 |
| Mild (AHI 5 to <15) | 4,252 (28.5%) | 413 (24.6%) | 20,562 (27.6%) | 1,319 (26.3%) |  |
| Moderate (AHI 15 to <30) | 3,952 (26.5%) | 456 (27.2%) | 20,316 (27.3%) | 1,418 (28.3%) |  |
| Severe (AHI ≥30) | 6,702 (45.0%) | 807 (48.2%) | 33,584 (45.1%) | 2,276 (45.4%) |  |
| **Comorbidity and context** |  |  |  |  |  |
| Elixhauser index | 4.1 ± 2.9 | 3.5 ± 2.5 | 4.8 ± 3.3 | 3.9 ± 2.8 | <0.001 |
| Charlson index | 1.4 ± 1.5 | 1.1 ± 1.3 | 1.8 ± 1.8 | 1.3 ± 1.5 | <0.001 |
| Hypertension | 7,669 (51.4%) | 783 (46.7%) | 43,964 (59.0%) | 2,663 (53.1%) | <0.001 |
| Depression | 6,626 (44.5%) | 672 (40.1%) | 32,652 (43.9%) | 1,982 (39.5%) | <0.001 |
| College Graduate (%) | 20.3 ± 10.3 | 19.0 ± 10.0 | 19.3 ± 10.1 | 18.4 ± 10.0 | <0.001 |
| Study-to-setup latency, d | 15 [9–28] | 11 [7–18] | 17 [9–32] | 11 [7–18] | <0.001 |
| **90-day CPAP Adherence** |  |  |  |  |  |
| CMS Adherence Rate | 6,840 (45.9%) | 661 (39.4%) | 32,782 (44.0%) | 1,753 (35.0%) | <0.001 |
| Mean hours/use | 3.5 ± 2.6 | 3.5 ± 2.5 | 3.5 ± 2.6 | 3.2 ± 2.6 | <0.001 |
| % used 4+ hours | 42.6 ± 34.9 | 33.9 ± 31.3 | 41.0 ± 34.9 | 30.6 ± 31.1 | <0.001 |

In the Questionnaire-Complete cohort, 45.9% met the 90-day CMS adherence definition. During days 331-360, 43.7% had no recorded PAP use (treatment abandonment), while 43.9% and 34.6% averaged at least 2 and at least 4 hours/night, respectively.

### Phase 1: Epidemiologic associations with Month-12 adherence

In the univariate demographic analysis **(Supplement S5)**, older age (odds ratio [OR] per SD, 1.11; 95% CI, 1.07-1.15) and male sex (vs female, 1.40; 1.31-1.51) were associated with greater odds of the primary adherence outcome. Significant differences in adherence were seen among the different races and ethnicities: compared with White patients, odds were lower among Asian (0.72; 0.65-0.81), Black (0.49; 0.41-0.57), Hispanic (0.60; 0.56-0.65), Other (0.69; 0.57-0.84), and Unknown (0.80; 0.66-0.97) categories.

In the primary multivariable analysis **(Figure 1)**, higher AHI (adjusted odds ratio [aOR] per SD, 1.36; 95% CI, 1.30-1.43) and longer self-reported sleep duration (per SD, 1.24; 1.20-1.29) were associated with greater odds of Month-12 mean use ≥4 hours/night. Higher Charlson comorbidity burden (per SD, 0.86; 0.83-0.90), depression (0.85; 0.78-0.92), and substance-use disorder (0.85; 0.78-0.93) were associated with lower odds. Study-to-setup latency was not associated with the primary outcome (per SD, 0.99; 0.96-1.03). Associations were broadly consistent for the secondary outcomes with the primary outcome **(Supplements S6-S8)**. Higher AHI, longer self-reported sleep duration, and snoring were positively associated with adherence or continued use, whereas claustrophobia, restless-legs symptoms, depression, and substance-use disorder were negatively associated.

**Figure 1.**
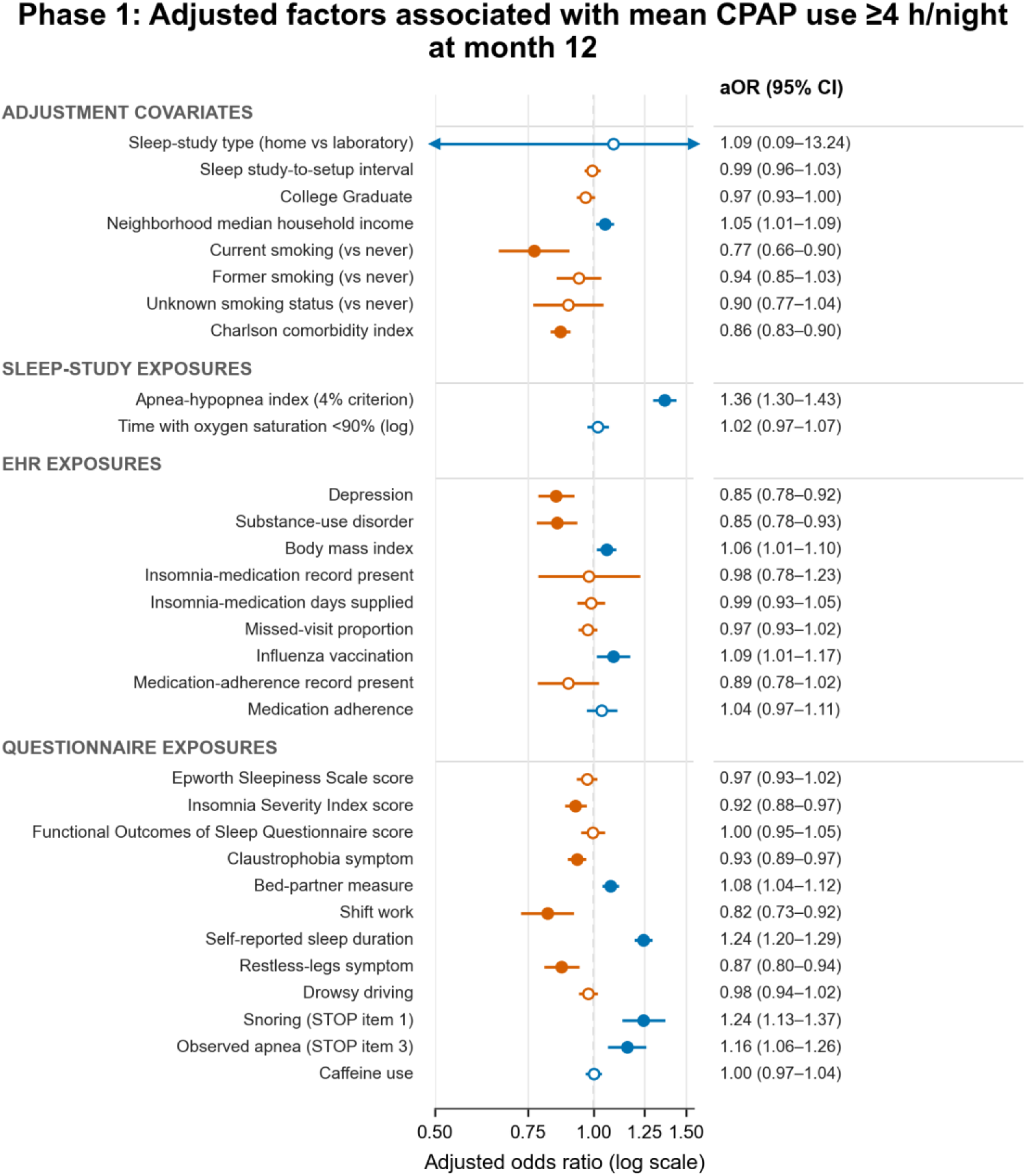
Phase 1 adjusted baseline associations with Month-12 adherence (mean use ≥4 hours/night during days 331-360). Continuous predictors are scaled per SD. Age, sex, and race and ethnicity remained adjustment covariates but are shown separately as univariate associations in Supplement S5. AHI indicates apnea–hypopnea index scored using a 4% desaturation criterion. Colored dots represent significant coefficients.

### Phase 2: Prediction of Month-12 primary and secondary adherence outcomes

For Phases 2-4, KPSC performance estimates in the main text and figures are reported for the random held-out test set. Prediction results are for XGBoost models unless specified otherwise. Findings remained stable in the temporal holdout set; detailed temporal validation results are provided in **Supplements S3 and S9-S12**.

For predicting the primary outcome (Month-12 mean use ≥4 hours/night), baseline-only AUROC was 0.678 (95% CI, 0.657-0.697). Across all four outcomes, baseline-only AUROCs ranged from 0.65 to 0.68 (logistic-regression models were developed only for baseline-only data and demonstrated comparable performance with generally slightly lower AUROCs ranging from 0.643 to 0.672; see **Supplement S13**). CPAP-data only discrimination for the primary outcome improved with each cumulative day of data **(Figure 2)**. AUROC increased from 0.660 after day 1 to 0.787 after day 7, 0.816 after day 14, and 0.854 after day 30. Adding baseline features (i.e., combined data models) incrementally improved performance through day 7, with the day-7 AUROC increasing from 0.787 to 0.810 (95% CI, 0.793–0.826). Secondary Month-12 outcomes for any use (abandonment) and mean use ≥2 hours/night showed similar performance patterns as for the primary outcome. Setup-date anchoring showed the same incremental pattern as with first-use anchoring but with lower overall discrimination **(Supplement S3)**.

**Figure 2.**
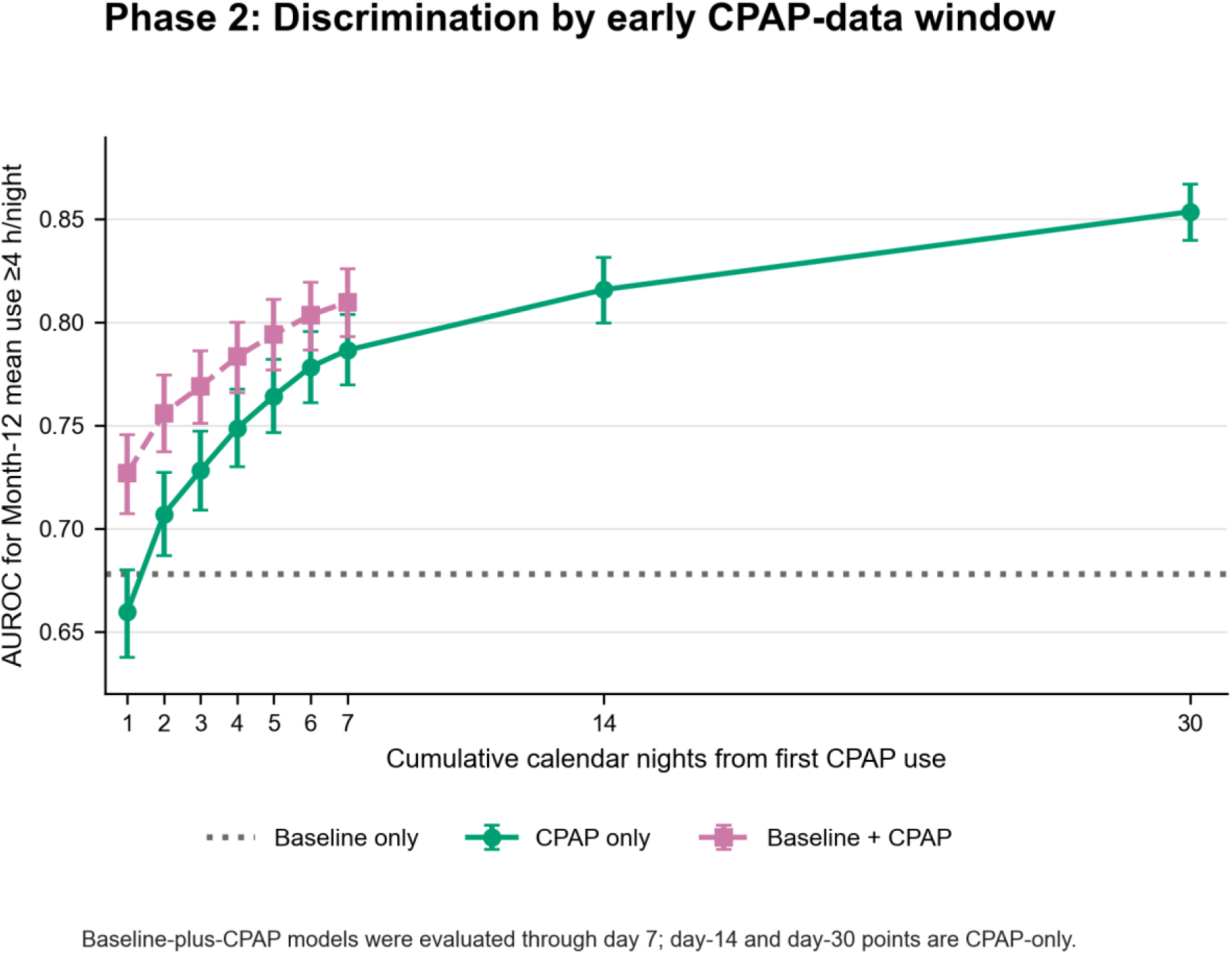
Phase 2 discrimination for the primary Month-12 outcome (mean use ≥4 hours/night during days 331– 360). AUROCs are reported for each cumulative nightly CPAP-data window beginning with the first positive-use night. The dotted line is baseline-only XGBoost; green and pink curves show CPAP-only and combined data XGBoost models. Error bars are bootstrap 95% confidence intervals. AUROC indicates area under the receiver-operating-characteristic curve.

In SHAP analyses of the primary outcome **(Figure 3)**, highest baseline attributions were sleep duration, AHI, race and ethnicity, sleep study analysis duration, age, and comorbidity, whereas CPAP data models were dominated by usage metrics with limited contributions from leak, pressure, and residual AHI parameters. Across all features in the baseline-only model, sleep study accounted for 28.0%, EHR 37.8%, and Questionnaire 34.2% of total mean absolute SHAP magnitude. In the combined data model, the corresponding shares were sleep-study 16.7%, EHR 23.6%, questionnaire 18.7%, and CPAP data 41.1%. Baseline model decision curves for all four outcomes reveal similar or higher net benefit for prediction models compared to “intervene on all” **(Supplement S14)**.

**Figure 3.**
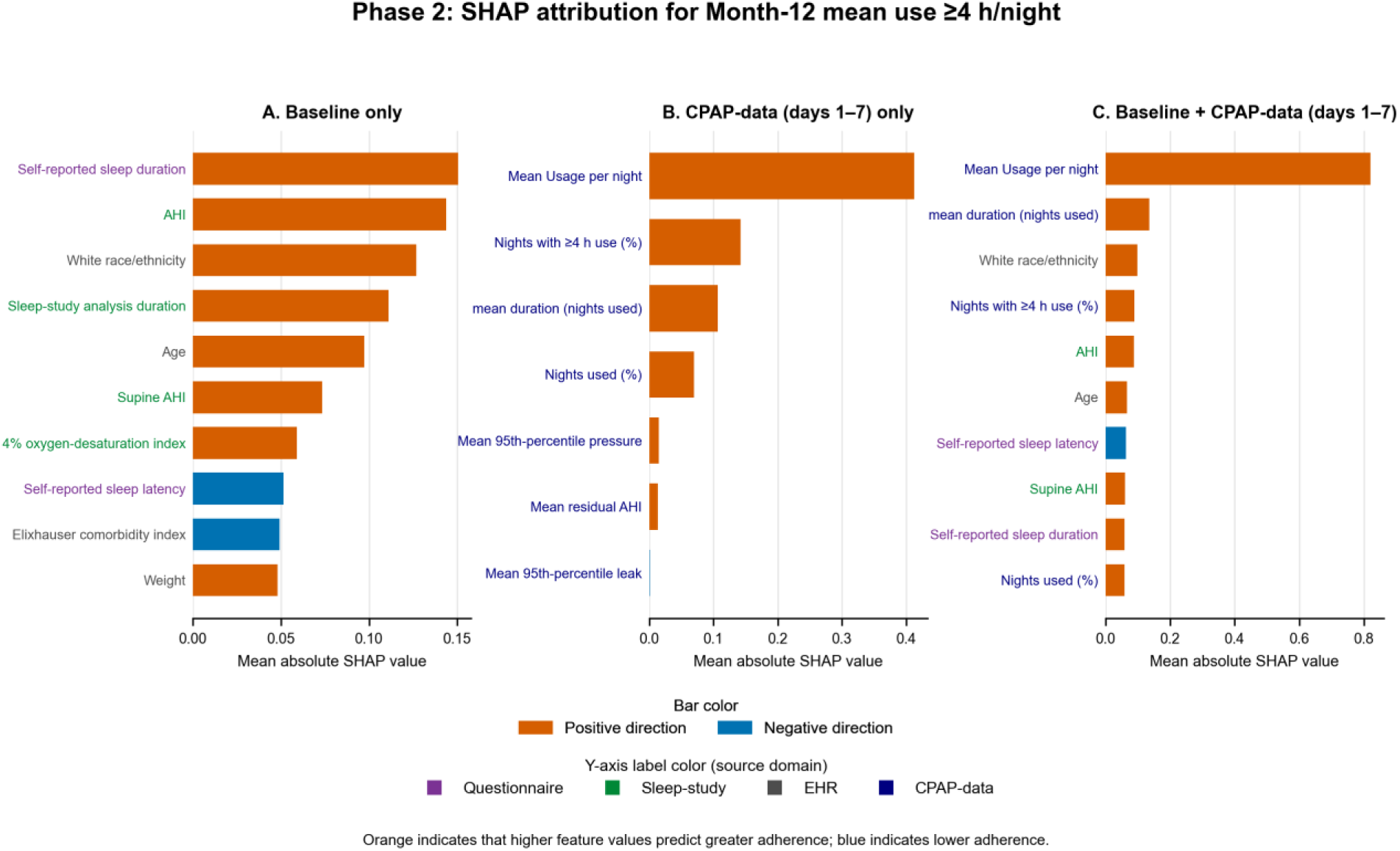
Phase 2 XGBoost feature attribution for the primary Month-12 outcome. Panels show (A) baseline-only, (B) CPAP-only data from days 1–7 only, and (C) combined data models. Bar length is mean absolute SHAP value. AHI indicates apnea–hypopnea index; SHAP, SHapley Additive exPlanations.

### Phase 3: Trajectory identification and prediction

Beginning on day 8, trajectories among 16,582 patients were Early-Decliners (7,116 [42.9%]), Stable-Moderate (3,147 [19.0%]), Gradual-Decliners (2,888 [17.4%]), and Stable-High (3,431 [20.7%]). The baseline and first-90-day CPAP characteristics of these trajectories are shown in **Supplement S15**. In the training cohort, nights used fell from 43.0% to 4.9% for Early-Decliners and from 83.0% to 25.7% for Gradual-Decliners, whereas Moderate and High Stable users remained consistent; mean duration on used nights changed from 3.3 to 3.7 and 5.3 to 4.4 hours/night, for the two stable groups respectively **(Figure 4)**. Assignment was reliable (entropy and minimum average posterior probability, both 0.989), and the selected four-class model was frozen before assignment in the validation cohorts.

**Figure 4.**
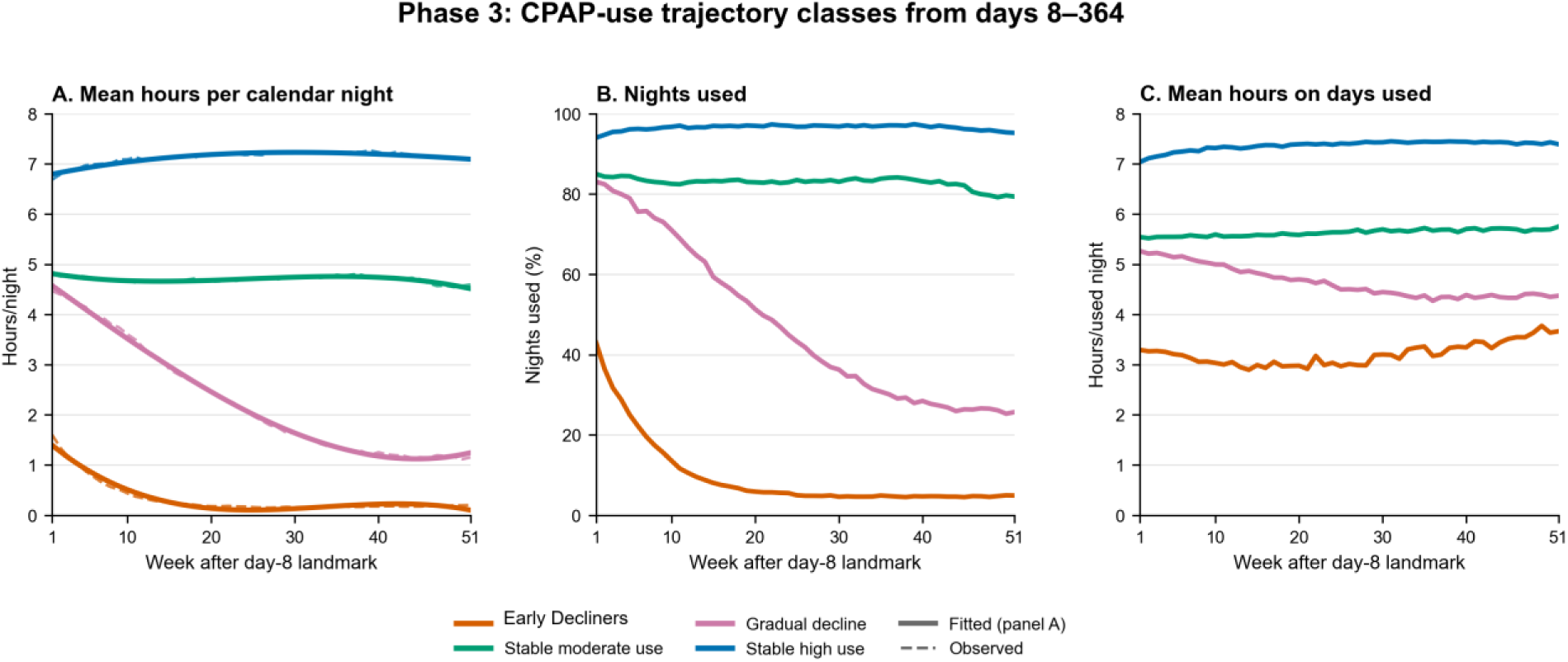
Phase 3 day-8-to-364 CPAP-use trajectory classes and observed component measures. (A) Observed and cubic Gaussian fitted weekly mean hours per calendar night used to define the classes, with nights without recorded use assigned zero. (B) Observed percentage of calendar nights with recorded use. (C) Observed mean hours per night among nights with recorded use. Panels B and C are descriptive summaries of the training-cohort classes defined in panel A, not additional trajectory-model outcomes. Calendar days 1-7 beginning with first positive use were excluded from the target and reserved for prediction. Classes are Early-Decliners (class 1), Stable-Moderate use (class 2), Gradual-Decliners (class 3), and Stable-High use (class 4).

Baseline features alone had limited discrimination to predict trajectories **(Figure 5 and Supplement S11)**: macro one-versus-rest AUROC was 0.619 (95% CI, 0.605-0.632). CPAP-only data from days 1-7 alone revealed higher macro AUROC to 0.756 (0.745-0.767), and combined data increased it further to 0.770 (0.759-0.781).

**Figure 5.**
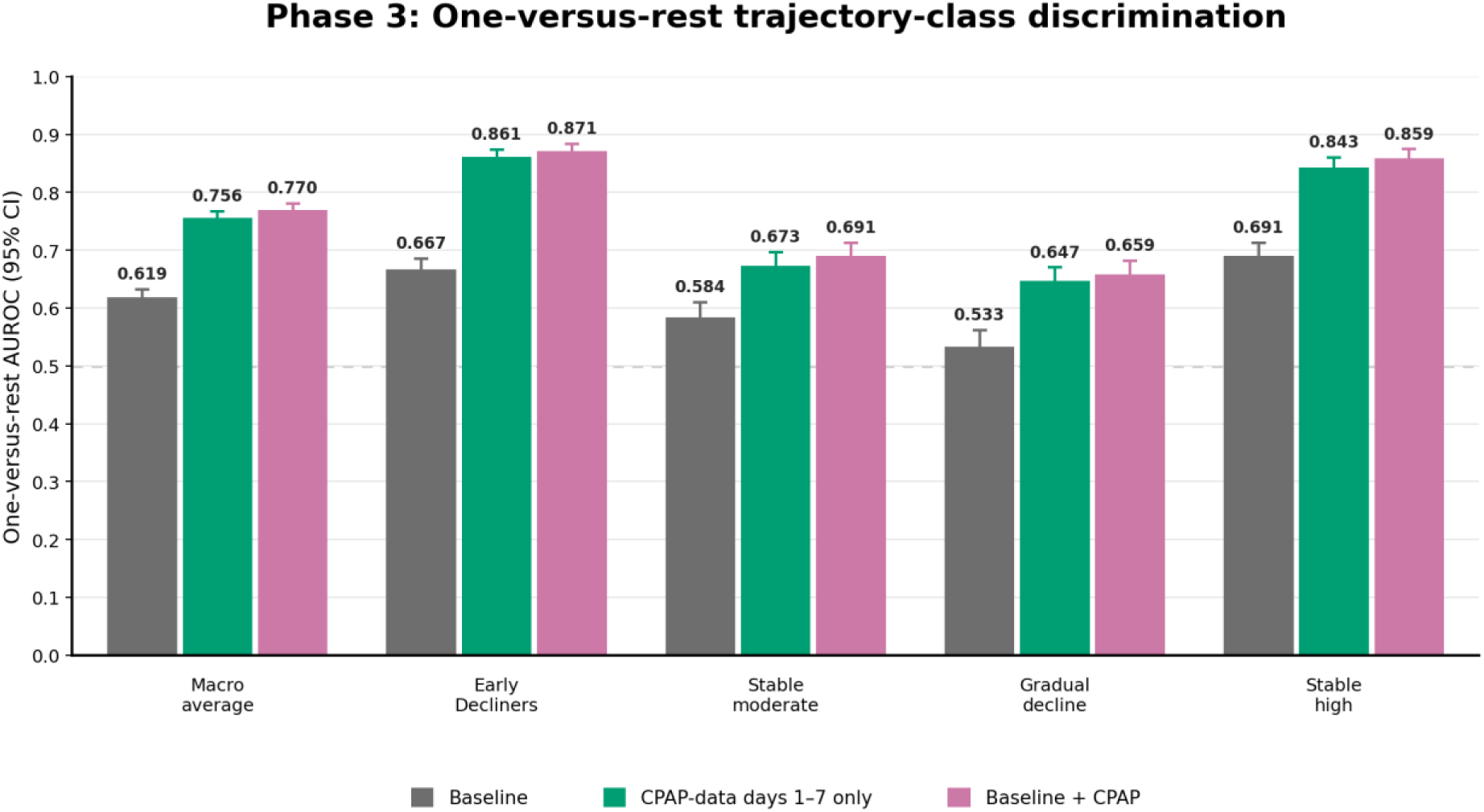
Phase 3 discrimination for prediction of day-8-to-364 CPAP trajectory class. Grouped vertical bars show macro-average and class-specific one-versus-rest AUROCs for baseline only, CPAP-data from days 1-7 only, and combined data models; error bars show bootstrap 95% confidence intervals.

For the combined model, class-specific AUROC was highest for Early-Decliners (0.871) and Stable-High (0.859) and lower for Stable-Moderate (0.691) and Gradual-Decliners (0.659). The CPAP-data only model showed the same pattern (Early-Decliners, 0.861; Stable-High users, 0.843; Stable-Moderate users, 0.673; Gradual-Decliners, 0.647). The full one-versus-rest results for all three models are reported in **Supplement S11**. Pairwise trajectory-class comparisons showed good-to-excellent discrimination for most contrasts involving Early-Decliners or Stable-High use; the combined model performed best for Early-Decliners versus Stable-High users (AUROC, 0.942; 95% CI, 0.931-0.952), whereas the principal difficulty was distinguishing Stable-Moderate users from Gradual-Decliners (AUROC, 0.581; 95% CI, 0.546-0.616; **Supplement S16**).

For baseline-only trajectory prediction, questionnaire features accounted for 38.9% of total mean absolute SHAP magnitude, EHR features for 30.9%, and sleep-study features for 30.2% **(Figure 6)**. The leading individual features were self-reported sleep duration, AHI, sleep-study analysis duration, White race and ethnicity, and age. After adding CPAP-data from days 1-7, CPAP-data features accounted for 48.0% of total SHAP magnitude; questionnaire, EHR, and sleep-study features accounted for 19.9%, 17.5%, and 14.6%, respectively. Mean hours per night, mean duration on used nights, percentage of nights with ≥4 hours of use, and percentage of nights used were the four leading combined-model features.

**Figure 6.**
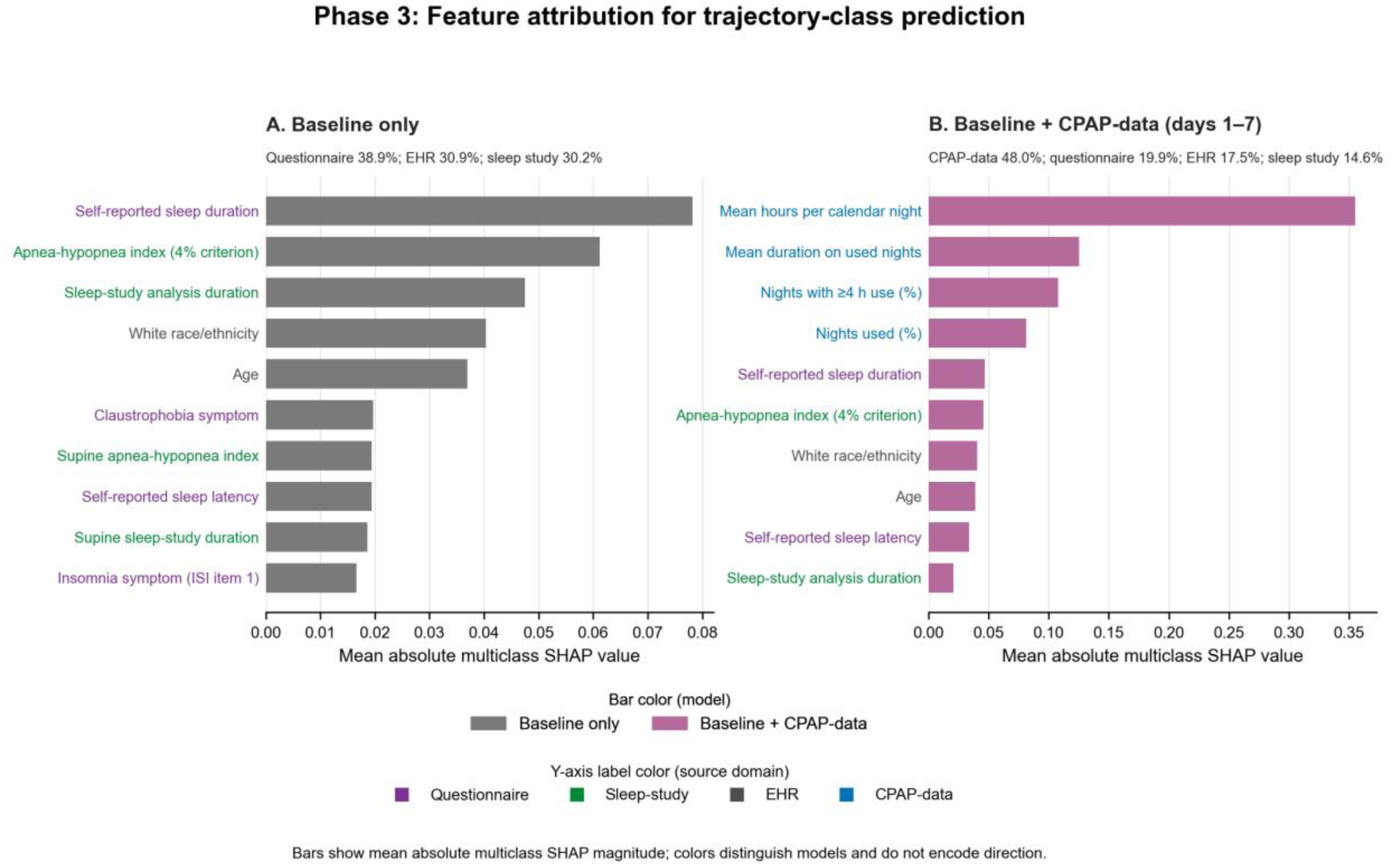
Phase 3 XGBoost feature attribution for prediction of day-8-to-364 CPAP trajectory class. Panels show (A) baseline-only and (B) combined baseline+CPAP (days 1-7) models. Bar length is the overall mean absolute multiclass SHAP value. SHAP indicates SHapley Additive exPlanations.

### Phase 4: Reduced-Feature Models and Implementation Considerations

In the KPSC held-out test set, the reduced-feature models retained discrimination comparable to the full models for the primary Phase 2 outcome (**Figure 7 and Supplement S17**). AUROCs for the full, parsimonious, and Questionnaire-Sleep-Study baseline models were 0.678, 0.644, and 0.672, respectively. For the combined data models, AUROCs were 0.811, 0.804, and 0.810 for the full, parsimonious, and Questionnaire-Sleep-Study models, respectively. The same pattern was observed for the secondary Month-12 and CMS outcomes and estimates with 95% confidence intervals are reported in **Supplement S17**.

**Figure 7.**
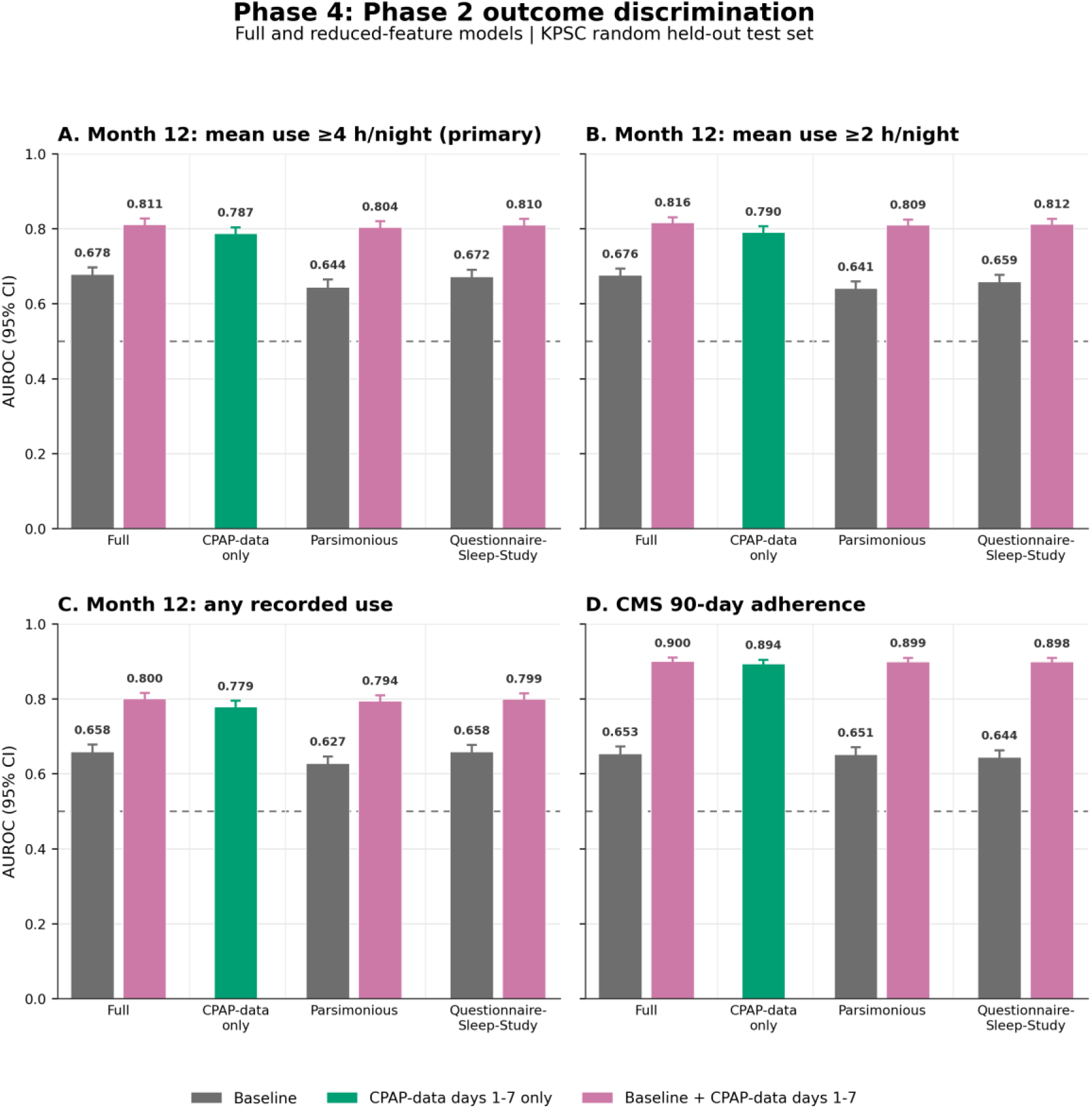
Phase 4 discrimination for Month-12 and CMS adherence outcomes across full and reduced-feature model specifications in the KPSC random held-out test set (n=2,G82). Panels show (A) the primary Month-12 outcome, mean CPAP use ≥4 hours/night; (B) Month-12 mean use ≥2 hours/night; (C) any Month-12 use; and (D) CMS 90-day adherence. Bars show AUROC and error bars show bootstrap 95% confidence intervals. Baseline models use pretreatment features; combined models add CPAP-data from days 1-7. The CPAP-data-only model is common across feature families. The post-treatment CMS models partially overlap the days 1-7 predictor window.

For trajectory prediction, macro-average AUROCs for the full, parsimonious, and Questionnaire-Sleep-Study baseline models were 0.619, 0.600, and 0.610, respectively. The CPAP-only model achieved an AUROC of 0.756 and for the combined data models, the corresponding AUROCs were 0.770, 0.767, and 0.768 (**Figure 8 and Supplement S18**). Across the combined models, discrimination remained highest for Early-Decliners and Stable-High and lower for Stable-Moderate and Gradual-Decliners.

**Figure 8.**
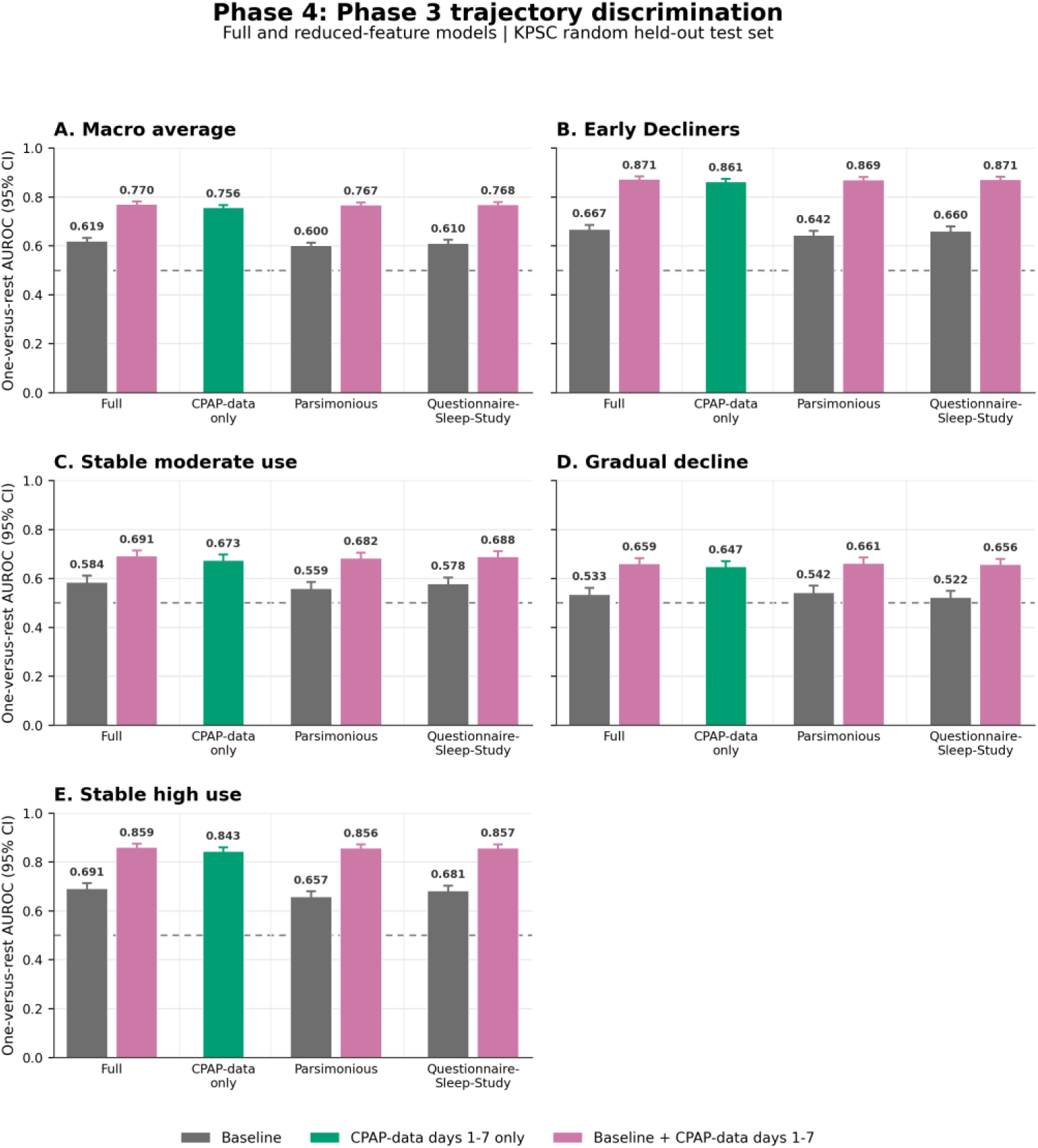
Phase 4 discrimination for prediction of day-8-to-364 CPAP trajectory class across full and reduced-feature model specifications in the KPSC random held-out test set (n=2,G81). Panels show (A) macro-average and class-specific one-versus-rest AUROC for (B) Early-Decliners, (C) Stable-Moderate users, (D) Gradual-Decliners, and (E) Stable-High users. Bars show AUROC and error bars show bootstrap 95% confidence intervals. Baseline models use pretreatment features and combined models add CPAP-data from days 1-7. The CPAP-data-only model is common across feature families.

## Discussion

This study aimed to evaluate predictors of long-term longitudinal CPAP adherence and revealed several key insights. First, multiple demographic, clinical, behavioral, and OSA-related characteristics derived from EHR, sleep studies, and questionnaires were independently associated with adherence at 1-year. Second, prediction models that utilized baseline pre-treatment features provided modest discrimination which substantially improved when incorporating early CPAP use (i.e., within the first 7 days). Usage metrics provided the primary signal without significant contributions from leak, pressure, nor residual AHI metrics. Third, longitudinal modeling identified four distinct trajectories of one-year use: Stable-High, Stable-Moderate, Gradual-Decliners, and Early-Decliners, and demonstrated that declining CPAP exposure was driven predominantly by fewer nights of use rather than progressively shorter use on nights when CPAP was applied. Early CPAP data similarly provided substantially stronger signals in predicting long-term trajectories compared to baseline-only data, especially for the Early-Decliners and High-Stable trajectories.

The epidemiologic analysis identified factors associated with long-term adherence consistent with those previously reported in literature, although often for short-term (i.e., 90-180 days) adherence. Greater OSA severity, older age, male sex, and socioeconomic affluence were associated with greater odds of adherence^34–36^, whereas greater comorbidity burden^36,37^ and depression^38^ were associated with lower odds. Substance-use disorder was also associated with lower adherence in our cohort, while prior studies have similarly linked alcohol use^39^, smoking^7,9^, and chronic opioid use^40^ with poorer CPAP adherence. We additionally analyzed EHR-derived behavioral patterns revealing that patients that received influenza vaccination within the past year were more likely to be adherent; on the other hand, insomnia medication use, appointment no-show rates, and hypertension and cholesterol medication adherence had no significant associations with CPAP adherence. For questionnaire-based information, longer self-reported sleep duration and existence of a bed partner predicted adherence while insomnia symptoms, shift work, claustrophobia, and restless legs predicted non-adherence. A prior study similarly associated insomnia symptoms with poorer CPAP adherence.^41^ In our study, ESS scores did not predict adherence, a factor that has variably been associated with adherence in literature.^42–46^ SHAP analysis of the XGBoost models identified adherence-related factors that were largely consistent with those in the primary logistic association model, with higher education additionally emerging as a predictor of better adherence.

The results of our prediction modeling extend the limited literature on CPAP adherence prediction. Pretreatment models have generally shown only modest discrimination, whereas models incorporating early CPAP use have performed better.^47–49^ Domínguez-Guerrero et al. reported an AUC of 0.71 using baseline pre-treatment features and increasing to 0.91 when incorporating the first 30-days of CPAP data when predicting one-year adherence. While these findings parallel our own results, our study further extends the evidence through a substantially larger and more diverse cohort, multiple early-use landmarks, and temporal validation. We also demonstrated that substantially improved predictive signal occurs early within the first week of CPAP use. Independent external validation, however, remains lacking across studies, including ours.

We explored whether our prediction models could have clinical utility. While decision curve analysis supports clinical utility, the overall performance using only baseline pre-treatment data was modest. Other factors such as treatment beliefs^10,50,51^, self-efficacy^50–52^, expectations, perceived benefit^10,51,52^, and physiologic endotypic factors^53–55^ (e.g., arousal threshold, airway collapsibility, loop gain) were unmeasured, which may have limited predictive performance. Models incorporating early CPAP use during the first week may be more useful given their greater predictive performances. Further, models predicting long-term trajectories performed particularly well in identifying Early-Decliners and High-Stable users, but had greater difficulty distinguishing Moderate-Stable from Gradual-Decliners, whose trajectories began with similar levels of CPAP use. These findings support the potential use of trajectory prediction to optimize follow-up workflows and guide resource allocation. For example, a patient predicted to be in the Early-Decliners trajectory may be prioritized for early intensive follow-up, while those in the Moderate-Stable or Gradual-Decliners may benefit from structured follow-up after 2-4 weeks and assess if declining usage is evident. Finally, SHAP values can provide guidance regarding the proper intervention. For example, concurrent cognitive behavioral therapy should be considered when the negative predictor relates to insomnia, shift-work, or claustrophobia. Nevertheless, we emphasize that these prediction models should not be used to determine eligibility for, restrict access to, or deny CPAP therapy which would create barriers to care and possibly exacerbate racial/ethnic disparities.

For implementation, XGBoost models, with recalibration, are likely preferred over the logistic regression approach due to the ability to inherently handle missingness and availability of SHAP values to guide personalized intervention.^56^ Model selection for implementation includes the following options: (1) CPAP-data only model which can incrementally update after each additional day between days 1-7 and possibly at 14 or 30 days; (2) Pre-treatment baseline models in which performance is modest but may be appropriately used to select patients for more intensive follow-up (but not to deny access to CPAP); (3) Combined models that incorporate both baseline and CPAP data post-treatment initiation, offering a more holistic approach (albeit with only incremental improved performance). If choosing to deploy models using baseline features, the *reduced-feature models* may be preferred for ease of implementation and mostly preserved performance, and the choice between the Parsimonious and Questionnaire-Sleep Study model may be guided by the platform hosting the models. For example, a middleware sleep management platform would be amenable to deploy the Questionnaire-Sleep Study model without need for EHR integration. Alternatively, if models are hosted in EHR with limited questionnaire information, the Parsimonious model may be preferred.

An important finding from this study is the racial and ethnic differences in adherence. Disparities among minoritized populations has been widely reported in literature.^5,8,57–59^ Compared with White patients, Asian, Black, and Hispanic patients had lower adjusted odds of long-term adherence despite adjustments for socioeconomic, clinical, OSA-related, and behavioral characteristics. This finding likely reflects unmeasured social, cultural, linguistic, environmental, and health-system factors, and highlights the need for tailoring care strategies to enhance personalized management and promote equitable outcomes. This may include the use of technology such as automated management to improve access to care and development of linguistically-tailored interventions.^4,60,61^

This study has several strengths. First, we evaluated long-term adherence while previous literature has predominantly reported short-term adherence (e.g., 90-180 days). Our evaluation was performed in a health system in which CPAP devices were purchased for patients at dispensation and not subject to retrieval if non-adherent. Second, our evaluation cohort was large with substantial demographic, clinical, and geographic diversity. Third, the study was performed in an integrated health system with automated data capture that enabled access to a broad set of EHR data, sleep study metrics, and patient-reported data from questionnaires. Fourth, we incorporated several EHR-derived behavioral measures that have been infrequently evaluated, including medication adherence, appointment no-show rates, influenza vaccination status, and insomnia medication days supplied (as a proxy for insomnia medication use frequency). Although we previously reported that several of these EHR-derived features were associated with adherence, in this larger analysis only influenza vaccination status remained a significant predictor.^62,63^

Several limitations should also be considered. First, this was an observational study within a single integrated health care system. Although KPSC is a network of 14 medical centers with independent sleep practices, similar clinical workflows could limit generalizability to other systems, especially since KPSC’s model of supporting access to CPAP equipment regardless of adherence is unusual in the United States. Second, despite the substantial size of our patient cohort, a minority had complete questionnaire data and therefore significantly reduced the primary analysis cohort. However, baseline characteristics were similar between the questionnaire-complete primary cohort and the larger EHR-Sleep-Study cohort which suggests ability to generalize insights from the smaller to larger cohort. Third, our questionnaire data was developed for clinical use and did not include important constructs such as treatment beliefs nor self-efficacy, important influences in CPAP adherence.^10,51,52,64–66^ Fourth, external validation was not performed and is required to demonstrate generalizability. Finally, a prospective study is necessary to truly validate clinical utility and to identify interventions that can modify adherence or trajectory outcomes.

In conclusion, CPAP adherence at one-year was associated with multiple demographic, clinical, behavioral, and OSA-related baseline factors. Although baseline-only prediction was modest, objective CPAP use during the first week provided substantially stronger prognostic information for both Month-12 adherence and long-term trajectory class, with baseline features adding incremental gain. These findings support potential clinical utility in guiding holistic and equitable CPAP management strategies to promote long-term adherence. External validation and prospective studies to evaluate implementation are needed.

## Supporting information

All Supplementals

## Data Availability

The individual-level data underlying this study are not publicly available because they contain protected health information. All aggregate results supporting the findings of this study are provided within the manuscript and its supplementary materials.

