## Supplementary material for "Predicting One-Year CPAP Adherence and Use Trajectories Utilizing Baseline and Early Use Data": All Supplementals

**Supplement S1.** Sleep-questionnaire questions and answer options.

**Supplement S2.** Predictor feature data dictionary.

**Supplement S3.** Phase 2 setup-date anchoring sensitivity analysis for the primary Month-12 outcome.

**Supplement S4.** Phase 4 reduced-feature model definitions.

**Supplement S5.** Phase 1 univariate demographic associations

**Supplement S6.** Phase 1 pre-initiation factors associated with CMS 90-day adherence.

**Supplement S7.** Phase 1 pre-initiation factors associated with Month-12 mean CPAP use ≥2 hours/night.

**Supplement S8.** Phase 1 pre-initiation factors associated with Month-12 treatment abandonment,

**Supplement S9.** Temporal calibration of Phase 2 XGBoost models for the primary Month-12 outcome.

**Supplement S10.** Temporal one-versus-rest calibration of Phase 3 XGBoost models for day-8-to-364 CPAP trajectory class.

**Supplement S11.** One-versus-rest AUROC for prediction of day-8-to-364 CPAP trajectory class.

**Supplement S12.** Temporal calibration measures for Phase 2 primary-outcome and Phase 3 trajectory-class prediction.

**Supplement S13.** Baseline-only logistic-regression and XGBoost AUROCs for the four Phase 2 outcomes.

**Supplement S14.** Baseline decision-curve analysis

**Supplement S15.** Baseline and first-90-day CPAP characteristics by day-8-to-364 CPAP-use trajectory class, with four-group omnibus comparisons.

**Supplement S16.** Conditional pairwise AUROC for prediction of day-8-to-364 CPAP trajectory class.

**Supplement S17.** Phase 4 held-out AUROC for Phase 2 outcomes across full and reduced-feature model specifications.

**Supplement S18.** Phase 4 held-out one-versus-rest AUROC for Phase 3 trajectory prediction across full and reduced-feature model specifications.

### Supplement S1. Sleep-questionnaire questions and answer options

The table reproduces all 81 ordered item rows from the supplied questionnaire workbook, including calculated scale-total rows.

| Item | Question | Response format, answer options, and notes |
|---|---|---|
| **General Sleep Information** | | |
| 1 | In a few words, describe your sleep problem(s) | **Response format:** Free text |
| 2 | Which of the following would you say is your chief complaint? | **Response format:** Single select<br>**Answer options:** Snoring / possible sleep apnea; Difficulty falling or staying asleep; Daytime sleepiness/fatigue; Abnormal behaviors/movements during sleep; Restless legs; Sleep schedule/circadian problem; PAP/sleep apnea treatment problem; Other |
| 3 | Which of these (if any) do you currently use during sleep? (select all that apply) | **Response format:** Multi-select<br>**Answer options:** None; CPAP/APAP; BiPAP/BPAP; Oral appliance; Supplemental oxygen; Hypoglossal nerve stimulator (e.g., Inspire); Positional therapy device; Other |
| 4 | Have you ever had any type of sleep study before? | **Response format:** Single select<br>**Answer options:** No; Yes; Not sure<br>**Scoring/notes:** *If Yes: Home sleep apnea test / In-lab sleep study / Both / Not sure* |
| 5 | How tall are you? | **Response format:** Numeric<br>**Answer options:** Feet + inches |
| 6 | How much do you weigh? | **Response format:** Numeric<br>**Answer options:** Pounds |
| 7 | What is your gender at birth? | **Response format:** Single select<br>**Answer options:** Male; Female |
| **Sleep and Wake Patterns** | | |
| 8 | What time do you get into bed? | **Response format:** Time |
| 9 | How long does it take you to fall asleep? | **Response format:** Numeric<br>**Answer options:** Minutes |
| 10 | How many times per night do you wake up? | **Response format:** Numeric<br>**Answer options:** Whole number |
| 11 | How long does it usually take to fall back asleep? | **Response format:** Numeric<br>**Answer options:** Minutes; N/A – I don't usually wake up |
| 12 | What is your usual wake time? | **Response format:** Time |
| 13 | How much sleep do you get on average each night? | **Response format:** Duration<br>**Answer options:** Hours + minutes |
| 14 | How many days per week do you take naps? | **Response format:** Numeric / single select<br>**Answer options:** 0; 1; 2; 3; 4; 5; 6; 7 days/week |
| 15 | How long are your naps on average? | **Response format:** Numeric<br>**Answer options:** Minutes; N/A – I don't nap |
| 16 | Do you work a night, split, or variable schedule shift? | **Response format:** Single select<br>**Answer options:** No; Yes – night shift; Yes – rotating/variable shift; Yes – split shift; Other |
| **Sleep-Disordered Breathing Risk (STOP-S)** | | |
| 17 | Do you snore loudly (louder than talking or loud enough to be heard through closed doors)? | **Response format:** Single select<br>**Answer options:** Yes; No |
| **Sleep-Disordered Breathing Risk (STOP-T)** | | |
| 18 | Do you often feel tired, fatigued, or sleepy during the daytime? | **Response format:** Single select<br>**Answer options:** Yes; No |

| Item | Question | Response format, answer options, and notes |
|---|---|---|
| **Sleep-Disordered Breathing Risk (STOP-O)** | | |
| 19 | Do you stop breathing during your sleep (or has anyone observed you do this)? | **Response format:** Single select<br>**Answer options:** Yes; No |
| **Sleep-Disordered Breathing Risk (STOP-P)** | | |
| 20 | Do you have or are you being treated for high blood pressure? | **Response format:** Single select<br>**Answer options:** Yes; No |
| **Sleep-Disordered Breathing Risk (STOP N)** | | |
| 21 | What is your collar size (or best guess if not known. Average is 16.0 inches [men] or 13.5 inches [women]) | **Response format:** Numeric<br>**Answer options:** Inches; Don't know |
| **Sleep-Disordered Breathing Risk** | | |
| 22 | How many days per week do you wake up with a headache? | **Response format:** Numeric / single select<br>**Answer options:** 0; 1; 2; 3; 4; 5; 6; 7 days/week |
| **Insomnia Severity Index** | | |
| 23 | Difficulty falling asleep | **Response format:** Single select<br>**Answer options:** 0 = None; 1 = Mild; 2 = Moderate; 3 = Severe; 4 = Very severe<br>**Scoring/notes:** *ISI item; score 0–4* |
| 24 | Difficulty staying asleep | **Response format:** Single select<br>**Answer options:** 0 = None; 1 = Mild; 2 = Moderate; 3 = Severe; 4 = Very severe<br>**Scoring/notes:** *ISI item; score 0–4* |
| 25 | Problems waking up too early | **Response format:** Single select<br>**Answer options:** 0 = None; 1 = Mild; 2 = Moderate; 3 = Severe; 4 = Very severe<br>**Scoring/notes:** *ISI item; score 0–4* |
| 26 | How satisfied/dissatisfied are you with your sleep pattern? | **Response format:** Single select<br>**Answer options:** 0 = Very satisfied; 1 = Satisfied; 2 = Moderately satisfied; 3 = Dissatisfied; 4 = Very dissatisfied<br>**Scoring/notes:** *ISI item; score 0–4* |
| 27 | How noticeable to others do you think your sleep problem is in terms of impairing the quality of your life? | **Response format:** Single select<br>**Answer options:** 0 = Not at all noticeable; 1 = A little; 2 = Somewhat; 3 = Much; 4 = Very much noticeable<br>**Scoring/notes:** *ISI item; score 0–4* |
| 28 | How worried/distressed are you about your sleep problem? | **Response format:** Single select<br>**Answer options:** 0 = Not at all worried; 1 = A little; 2 = Somewhat; 3 = Much; 4 = Very much worried<br>**Scoring/notes:** *ISI item; score 0–4* |
| 29 | To what extent does your sleep problem interfere with daily functioning? | **Response format:** Single select<br>**Answer options:** 0 = Not at all interfering; 1 = A little; 2 = Somewhat; 3 = Much; 4 = Very much interfering<br>**Scoring/notes:** *ISI item; score 0–4* |
| **Additional Sleep Questions** | | |
| 30 | How often do you take medication to help you sleep? | **Response format:** Single select<br>**Answer options:** Never; Less than once per week; 1–2 nights/week; 3–4 nights/week; 5–7 nights/week |
| 31 | How many days per week are you NOT satisfied with your sleep? | **Response format:** Numeric / single select<br>**Answer options:** 0; 1; 2; 3; 4; 5; 6; 7 days/week |
| 32 | How many days per week is sleepiness a problem? | **Response format:** Numeric / single select<br>**Answer options:** 0; 1; 2; 3; 4; 5; 6; 7 days/week |
| 33 | How many times per DAY do you have a caffeinated drink? | **Response format:** Single select / numeric<br>**Answer options:** 0; 1; 2; 3; 4; 5; 6+ |
| 34 | How often do you feel sleepy or tired during your waking time? | **Response format:** Single select<br>**Answer options:** Never; Rarely; Sometimes; Often; Almost always |

| Item | Question | Response format, answer options, and notes |
|---|---|---|
| 35 | How often do you nod off or fall asleep while driving? | **Response format:** Single select<br>**Answer options:** Never; Rarely; Sometimes; Often; Almost always; I do not drive |
| 36 | How often do you experience claustrophobia or anxiety about confined spaces? | **Response format:** Single select<br>**Answer options:** Never; Rarely; Sometimes; Often; Almost always |
| 37 | How often do you sleep in the same room as another person? | **Response format:** Single select<br>**Answer options:** Never; Rarely; Sometimes; Often; Almost always |
| 38 | How often do you have unusual movements during sleep (e.g., kicking, punching, or sleep-walking)? | **Response format:** Single select<br>**Answer options:** Never; Rarely; Sometimes; Often; Almost always; Don't know |
| **Narcolepsy-Related Symptoms / PTSD** | | |
| 39 | Do you regularly experience 'sleep attacks' (falling asleep suddenly without meaning to)? | **Response format:** Single select<br>**Answer options:** Yes; No |
| 40 | If yes, how often do sleep attacks occur? | **Response format:** Single select<br>**Answer options:** Less than once/month; 1–3 times/month; 1–2 times/week; 3–6 times/week; Daily or more |
| 41 | Have you EVER experienced 'sleep paralysis' (feel paralyzed on waking or falling asleep)? | **Response format:** Single select<br>**Answer options:** Yes; No |
| 42 | If yes, how often does sleep paralysis occur? | **Response format:** Single select<br>**Answer options:** Less than once/month; 1–3 times/month; 1–2 times/week; 3–6 times/week; Daily or more |
| 43 | Have you EVER experienced 'sleep hallucinations' (see, hear, or feel things that aren't real on waking or falling asleep)? | **Response format:** Single select<br>**Answer options:** Yes; No |
| 44 | If yes, how often do sleep hallucinations occur? | **Response format:** Single select<br>**Answer options:** Less than once/month; 1–3 times/month; 1–2 times/week; 3–6 times/week; Daily or more |
| 45 | Have you EVER experienced 'cataplexy' (SUDDEN muscle weakness when you laugh or feel strong emotion)? | **Response format:** Single select<br>**Answer options:** Yes; No |
| 46 | If yes, how often does cataplexy occur? | **Response format:** Single select<br>**Answer options:** Less than once/month; 1–3 times/month; 1–2 times/week; 3–6 times/week; Daily or more |
| 47 | Do you or your doctor think you may have post-traumatic stress disorder (PTSD)? | **Response format:** Single select<br>**Answer options:** No; Yes |
| **Epworth Sleepiness Scale** | | |
| 48a | Sitting and reading | **Response format:** Single select<br>**Answer options:** 0 = Would never doze; 1 = Slight chance of dozing; 2 = Moderate chance of dozing; 3 = High chance of dozing<br>**Scoring/notes:** *ESS item; score 0–3* |
| 48b | Watching television | **Response format:** Single select<br>**Answer options:** 0 = Would never doze; 1 = Slight chance of dozing; 2 = Moderate chance of dozing; 3 = High chance of dozing<br>**Scoring/notes:** *ESS item; score 0–3* |
| 48c | Sitting, inactive, in a public place (e.g., in a meeting, theater, or dinner event) | **Response format:** Single select<br>**Answer options:** 0 = Would never doze; 1 = Slight chance of dozing; 2 = Moderate chance of dozing; 3 = High chance of dozing<br>**Scoring/notes:** *ESS item; score 0–3* |
| 48d | As a passenger in a car for an hour or more without stopping for a break | **Response format:** Single select<br>**Answer options:** 0 = Would never doze; 1 = Slight chance of dozing; 2 = Moderate chance of dozing; 3 = High chance of dozing<br>**Scoring/notes:** *ESS item; score 0–3* |

| Item | Question | Response format, answer options, and notes |
|---|---|---|
| 48e | Lying down to rest when circumstances permit | **Response format:** Single select<br>**Answer options:** 0 = Would never doze; 1 = Slight chance of dozing; 2 = Moderate chance of dozing; 3 = High chance of dozing<br>**Scoring/notes:** *ESS item; score 0–3* |
| 48f | Sitting and talking to someone | **Response format:** Single select<br>**Answer options:** 0 = Would never doze; 1 = Slight chance of dozing; 2 = Moderate chance of dozing; 3 = High chance of dozing<br>**Scoring/notes:** *ESS item; score 0–3* |
| 48g | Sitting quietly after a meal without alcohol | **Response format:** Single select<br>**Answer options:** 0 = Would never doze; 1 = Slight chance of dozing; 2 = Moderate chance of dozing; 3 = High chance of dozing<br>**Scoring/notes:** *ESS item; score 0–3* |
| 48h | In a car, while stopped for a few minutes in traffic or at a light | **Response format:** Single select<br>**Answer options:** 0 = Would never doze; 1 = Slight chance of dozing; 2 = Moderate chance of dozing; 3 = High chance of dozing<br>**Scoring/notes:** *ESS item; score 0–3* |
| 48 Total | *Epworth Sleepiness Scale composite score* | **Response format:** Calculated<br>**Scoring/notes:** *Sum 48a–48h; range 0–24* |
| **FOSQ10** | | |
| 49a | Concentrating on the things you do | **Response format:** Single select<br>**Answer options:** 0 = Don't do this activity for other reasons / N/A; 1 = Yes, extreme difficulty; 2 = Yes, moderate difficulty; 3 = Yes, a little difficulty; 4 = No difficulty<br>**Scoring/notes:** *FOSQ-10 item; 0 denotes N/A/non-applicable activity* |
| 49b | Remembering things | **Response format:** Single select<br>**Answer options:** 0 = Don't do this activity for other reasons / N/A; 1 = Yes, extreme difficulty; 2 = Yes, moderate difficulty; 3 = Yes, a little difficulty; 4 = No difficulty<br>**Scoring/notes:** *FOSQ-10 item; 0 denotes N/A/non-applicable activity* |
| 49c | Operating a motor vehicle for SHORT distances (less than 100 miles) | **Response format:** Single select<br>**Answer options:** 0 = Don't do this activity for other reasons / N/A; 1 = Yes, extreme difficulty; 2 = Yes, moderate difficulty; 3 = Yes, a little difficulty; 4 = No difficulty<br>**Scoring/notes:** *FOSQ-10 item; 0 denotes N/A/non-applicable activity* |
| 49d | Operating a motor vehicle for LONG distances (more than 100 miles) | **Response format:** Single select<br>**Answer options:** 0 = Don't do this activity for other reasons / N/A; 1 = Yes, extreme difficulty; 2 = Yes, moderate difficulty; 3 = Yes, a little difficulty; 4 = No difficulty<br>**Scoring/notes:** *FOSQ-10 item; 0 denotes N/A/non-applicable activity* |
| 49e | Visiting your family or friends in their home | **Response format:** Single select<br>**Answer options:** 0 = Don't do this activity for other reasons / N/A; 1 = Yes, extreme difficulty; 2 = Yes, moderate difficulty; 3 = Yes, a little difficulty; 4 = No difficulty<br>**Scoring/notes:** *FOSQ-10 item; 0 denotes N/A/non-applicable activity* |
| 49f | Watching a movie or video | **Response format:** Single select<br>**Answer options:** 0 = Don't do this activity for other reasons / N/A; 1 = Yes, extreme difficulty; 2 = Yes, moderate difficulty; 3 = Yes, a little difficulty; 4 = No difficulty<br>**Scoring/notes:** *FOSQ-10 item; 0 denotes N/A/non-applicable activity* |
| 49g | Being as active as you want to be in the MORNING | **Response format:** Single select<br>**Answer options:** 0 = Don't do this activity for other reasons / N/A; 1 = Yes, extreme difficulty; 2 = Yes, moderate difficulty; 3 = Yes, a little difficulty; 4 = No difficulty<br>**Scoring/notes:** *FOSQ-10 item; 0 denotes N/A/non-applicable activity* |

| Item | Question | Response format, answer options, and notes |
|---|---|---|
| 49h | Being as active as you want to be in the EVENING | **Response format:** Single select<br>**Answer options:** 0 = Don't do this activity for other reasons / N/A; 1 = Yes, extreme difficulty; 2 = Yes, moderate difficulty; 3 = Yes, a little difficulty; 4 = No difficulty<br>**Scoring/notes:** *FOSQ-10 item; 0 denotes N/A/non-applicable activity* |
| 49i | Your relationship with family, friends or work colleagues | **Response format:** Single select<br>**Answer options:** 0 = Don't do this activity for other reasons / N/A; 1 = Yes, extreme difficulty; 2 = Yes, moderate difficulty; 3 = Yes, a little difficulty; 4 = No difficulty<br>**Scoring/notes:** *FOSQ-10 item; 0 denotes N/A/non-applicable activity* |
| 49j | Your mood | **Response format:** Single select<br>**Answer options:** 0 = Don't do this activity for other reasons / N/A; 1 = Yes, extreme difficulty; 2 = Yes, moderate difficulty; 3 = Yes, a little difficulty; 4 = No difficulty<br>**Scoring/notes:** *FOSQ-10 item; 0 denotes N/A/non-applicable activity* |
| 49 Total | *FOSQ10 composite score* | **Response format:** Calculated<br>**Scoring/notes:** *Conventionally expressed on a 5–20 scale; higher = better functional status* |
| **PHQ2** | | |
| 50a | Little interest or pleasure in doing things | **Response format:** Single select<br>**Answer options:** 0 = Not at all; 1 = Several days; 2 = More than half the days; 3 = Nearly every day<br>**Scoring/notes:** *Reflects last 2 weeks; score 0–3* |
| 50b | Feeling down, depressed, or hopeless | **Response format:** Single select<br>**Answer options:** 0 = Not at all; 1 = Several days; 2 = More than half the days; 3 = Nearly every day<br>**Scoring/notes:** *Reflects last 2 weeks; score 0–3* |
| 50 Total | *PHQ2 composite score* | **Response format:** Calculated<br>**Scoring/notes:** *Sum 50a–50b; range 0–6* |
| **Restless Legs Symptoms** | | |
| 51 | Do you experience unpleasant sensations in your legs combined with an urge to move them? | **Response format:** Single select<br>**Answer options:** Yes; No |
| 52 | Do these sensations occur mainly at rest and improve with movement? | **Response format:** Single select<br>**Answer options:** Yes; No; N/A |
| 53 | Are these sensations worse in the evening (compared to the morning)? | **Response format:** Single select<br>**Answer options:** Yes; No; N/A |
| 54 | Does it usually occur in the legs, arms, or both? | **Response format:** Single select<br>**Answer options:** Legs only; Arms only; Both legs and arms; N/A |
| 55 | If yes, how often do these sensations in your legs/arms occur? | **Response format:** Single select<br>**Answer options:** Never; Rarely (≤1 time/month); Sometimes (2–4 times/month); Often (5–15 times/month); Very often (≥16 times/month) |
| **Background** | | |
| 56 | What is your CURRENT employment status? | **Response format:** Single select<br>**Answer options:** Employed full-time; Employed part-time; Self-employed; Student; Homemaker or caregiver; Unemployed; Retired; Unable to work or disabled; Other; Prefer not to answer |
| 57 | What is your highest degree of schooling completed? | **Response format:** Single select<br>**Answer options:** Less than high school; High school or GED; Some college; Associate or technical degree; Bachelor's degree; Graduate or professional degree; Other; Prefer not to answer |
| 58 | What is your marital status? | **Response format:** Single select<br>**Answer options:** Never married/single; Married; Domestic partnership; Separated; Divorced; Widowed; Prefer not to answer |

| Item | Question | Response format, answer options, and notes |
|---|---|---|
| 59 | Are you Hispanic, Latino, or of Spanish origin? | **Response format:** Single select<br>**Answer options:** Yes; No; Prefer not to answer |
| 60 | What is your race (select ALL that apply)? | **Response format:** Multi-select<br>**Answer options:** American Indian or Alaska Native; Asian; Black or African American; Native Hawaiian or Other Pacific Islander; White; Other; Prefer not to answer |
| 61 | Do you have a commercial driver's license? | **Response format:** Single select<br>**Answer options:** Yes; No |

### Supplement S2. Predictor feature data dictionary

The table lists the raw predictor fields used in the reported model specifications. Source domains are EHR, Sleep Study, Questionnaire, and CPAP. Model-membership abbreviations are: QC baseline=Questionnaire-Complete baseline model; EHR baseline=EHR-Sleep-Study baseline model; Baseline + CPAP=combined baseline and early-CPAP model; and CPAP only=early-CPAP model.

| Raw feature | Plain-language definition | Source | Units/coding | Model membership* |
|---|---|---|---|---|
| **EHR** | | | | |
| icdx_e_ch_mi | Charlson component: Myocardial infarction | EHR | binary indicator | QC baseline; EHR baseline; Baseline + CPAP |
| icdx_e_ch_chf | Charlson component: Congestive heart failure | EHR | binary indicator | QC baseline; EHR baseline; Baseline + CPAP |
| icdx_e_ch_pvd | Charlson component: Peripheral vascular disease | EHR | binary indicator | QC baseline; EHR baseline; Baseline + CPAP |
| icdx_e_ch_cevd | Charlson component: Cerebrovascular disease | EHR | binary indicator | QC baseline; EHR baseline; Baseline + CPAP |
| icdx_e_ch_dem | Charlson component: Dementia | EHR | binary indicator | QC baseline; EHR baseline; Baseline + CPAP |
| icdx_e_ch_copd | Charlson component: Chronic pulmonary disease | EHR | binary indicator | QC baseline; EHR baseline; Baseline + CPAP |
| icdx_e_ch_rheum | Charlson component: Rheumatic/connective-tissue disease | EHR | binary indicator | QC baseline; EHR baseline; Baseline + CPAP |
| icdx_e_ch_pud | Charlson component: Peptic ulcer disease | EHR | binary indicator | QC baseline; EHR baseline; Baseline + CPAP |
| icdx_e_ch_mildld | Charlson component: Mild liver disease | EHR | binary indicator | QC baseline; EHR baseline; Baseline + CPAP |
| icdx_e_ch_diab_nc | Charlson component: Diabetes without chronic complications | EHR | binary indicator | QC baseline; EHR baseline; Baseline + CPAP |
| icdx_e_ch_diab_c | Charlson component: Diabetes with chronic complications | EHR | binary indicator | QC baseline; EHR baseline; Baseline + CPAP |
| icdx_e_ch_para | Charlson component: Hemiplegia or paraplegia | EHR | binary indicator | QC baseline; EHR baseline; Baseline + CPAP |
| icdx_e_ch_rd | Charlson component: Renal disease | EHR | binary indicator | QC baseline; EHR baseline; Baseline + CPAP |
| icdx_e_ch_cancer | Charlson component: Malignancy | EHR | binary indicator | QC baseline; EHR baseline; Baseline + CPAP |
| icdx_e_ch_msld | Charlson component: Moderate or severe liver disease | EHR | binary indicator | QC baseline; EHR baseline; Baseline + CPAP |
| icdx_e_ch_mets | Charlson component: Metastatic solid tumor | EHR | binary indicator | QC baseline; EHR baseline; Baseline + CPAP |
| icdx_e_ch_hiv | Charlson component: HIV/AIDS | EHR | binary indicator | QC baseline; EHR baseline; Baseline + CPAP |
| Charlson | Charlson comorbidity index | EHR | Index points | QC baseline; EHR baseline; Baseline + CPAP |
| icdx_e_ex_Arrhy | Elixhauser component: Cardiac arrhythmia | EHR | binary indicator | QC baseline; EHR baseline; Baseline + CPAP |
| icdx_e_ex_VD | Elixhauser component: Valvular disease | EHR | binary indicator | QC baseline; EHR baseline; Baseline + CPAP |
| icdx_e_ex_PCD | Elixhauser component: Pulmonary circulation disorder | EHR | binary indicator | QC baseline; EHR baseline; Baseline + CPAP |
| icdx_e_ex_HPTN_NC | Elixhauser component: Uncomplicated hypertension | EHR | binary indicator | QC baseline; EHR baseline; Baseline + CPAP |
| icdx_e_ex_HPTN_C | Elixhauser component: Complicated hypertension | EHR | binary indicator | QC baseline; EHR baseline; Baseline + CPAP |
| icdx_e_ex_OthND | Elixhauser component: Other neurologic disorder | EHR | binary indicator | QC baseline; EHR baseline; Baseline + CPAP |
| icdx_e_ex_Hptothy | Elixhauser component: Hypothyroidism | EHR | binary indicator | QC baseline; EHR baseline; Baseline + CPAP |
| icdx_e_ex_LD | Elixhauser component: Liver disease | EHR | binary indicator | QC baseline; EHR baseline; Baseline + CPAP |

| Raw feature | Plain-language definition | Source | Units/coding | Model membership* |
|---|---|---|---|---|
| icdx_e_ex_Lymp | Elixhauser component: Lymphoma | EHR | binary indicator | QC baseline; EHR baseline; Baseline + CPAP |
| icdx_e_ex_Tumor | Elixhauser component: Solid tumor without metastasis | EHR | binary indicator | QC baseline; EHR baseline; Baseline + CPAP |
| icdx_e_ex_Coag | Elixhauser component: Coagulopathy | EHR | binary indicator | QC baseline; EHR baseline; Baseline + CPAP |
| icdx_e_ex_Obesity | Elixhauser component: Obesity | EHR | binary indicator | QC baseline; EHR baseline; Baseline + CPAP |
| icdx_e_ex_WL | Elixhauser component: Weight loss | EHR | binary indicator | QC baseline; EHR baseline; Baseline + CPAP |
| icdx_e_ex_Fluid | Elixhauser component: Fluid or electrolyte disorder | EHR | binary indicator | QC baseline; EHR baseline; Baseline + CPAP |
| icdx_e_ex_BLA | Elixhauser component: Blood-loss anemia | EHR | binary indicator | QC baseline; EHR baseline; Baseline + CPAP |
| icdx_e_ex_DA | Elixhauser component: Deficiency anemia | EHR | binary indicator | QC baseline; EHR baseline; Baseline + CPAP |
| icdx_e_ex_Alcohol | Elixhauser component: Alcohol-use disorder | EHR | binary indicator | QC baseline; EHR baseline; Baseline + CPAP |
| icdx_e_ex_Drug | Elixhauser component: Drug-use disorder | EHR | binary indicator | QC baseline; EHR baseline; Baseline + CPAP |
| icdx_e_ex_Psycho | Elixhauser component: Psychosis | EHR | binary indicator | QC baseline; EHR baseline; Baseline + CPAP |
| icdx_e_ex_Dep | Elixhauser component: Depression | EHR | binary indicator | QC baseline; EHR baseline; Baseline + CPAP |
| Elixhauser | Elixhauser comorbidity index | EHR | Index points | QC baseline; EHR baseline; Baseline + CPAP |
| HT | Most recent pre-index height | EHR | inches | QC baseline; EHR baseline; Baseline + CPAP |
| WT | Most recent pre-index weight | EHR | pounds | QC baseline; EHR baseline; Baseline + CPAP |
| BMI | Body mass index | EHR | $kg/m^2$ | QC baseline; EHR baseline; Baseline + CPAP |
| DBP | Diastolic blood pressure | EHR | mmHg | QC baseline; EHR baseline; Baseline + CPAP |
| SBP | Systolic blood pressure | EHR | mmHg | QC baseline; EHR baseline; Baseline + CPAP |
| TEMPERATURE | Body temperature | EHR | °F | QC baseline; EHR baseline; Baseline + CPAP |
| PULSE | Clinical pulse rate | EHR | beats/min | QC baseline; EHR baseline; Baseline + CPAP |
| office_visit | Pre-index office-visit count | EHR | count | QC baseline; EHR baseline; Baseline + CPAP |
| GENDER | Recorded sex/gender used by the model | EHR | category; F=0, M=1 | QC baseline; EHR baseline; Baseline + CPAP |
| RACE_ETH | Race/ethnicity category | EHR | nominal category; one-hot encoded | QC baseline; EHR baseline; Baseline + CPAP |
| age | Age at index sleep study | EHR | years | QC baseline; EHR baseline; Baseline + CPAP |
| EDUCATION3 | Area-level educational-attainment measure 3 | EHR | percentage points | QC baseline; EHR baseline; Baseline + CPAP |
| College Graduate | Area-level percentage of college graduates | EHR | percentage points | QC baseline; EHR baseline; Baseline + CPAP |
| h_med_income_gp | Neighborhood median household-income group | EHR | ordinal category; 0/1/2 | QC baseline; EHR baseline; Baseline + CPAP |
| flu | Influenza-vaccination record | EHR | indicator | QC baseline; EHR baseline; Baseline + CPAP |
| no_show | Missed-appointment count | EHR | count | QC baseline; EHR baseline; Baseline + CPAP |
| no_show_pct | Missed-appointment ratio as stored | EHR | unitless ratio | QC baseline; EHR baseline; Baseline + CPAP |

| Raw feature | Plain-language definition | Source | Units/coding | Model membership* |
|---|---|---|---|---|
| ANTIDEPRESSANTS | Pre-index antidepressant exposure | EHR | medication/fill count | QC baseline; EHR baseline; Baseline + CPAP |
| med_adherence | Mean chronic-medication proportion of days covered | EHR | fraction | QC baseline; EHR baseline; Baseline + CPAP |
| insomnia_days_supply | Pre-index insomnia-medication days supplied | EHR | days | QC baseline; EHR baseline; Baseline + CPAP |
| insomnia_med_count | Number of insomnia-medication classes/records | EHR | count | QC baseline; EHR baseline; Baseline + CPAP |
| SMOKING_STATUS | Smoking-status category | EHR | nominal category; one-hot encoded | QC baseline; EHR baseline; Baseline + CPAP |
| setup_latency | Interval from index sleep study to CPAP setup | EHR | days | QC baseline; EHR baseline; Baseline + CPAP |
| **Sleep study** | | | | |
| HSAT/PSG | Diagnostic sleep-study type | Sleep study | category; HSAT=0, PSG=1 | QC baseline; EHR baseline; Baseline + CPAP |
| ANALYSIS_DURATION | Sleep-study analysis duration | Sleep study | minutes | QC baseline; EHR baseline; Baseline + CPAP |
| SUPINE_DURATION | Supine sleep-study duration | Sleep study | minutes | QC baseline; EHR baseline; Baseline + CPAP |
| SUPINE_DURATION_ PERCENT | Fraction of study spent supine | Sleep study | fraction | QC baseline; EHR baseline; Baseline + CPAP |
| ODI4PCT | 4% oxygen-desaturation index | Sleep study | events/hour | QC baseline; EHR baseline; Baseline + CPAP |
| ODT4PCT_SUPINE | Supine 4% oxygen-desaturation index | Sleep study | events/hour | QC baseline; EHR baseline; Baseline + CPAP |
| ODI4PCT_NONSUPINE | Non-supine 4% oxygen-desaturation index | Sleep study | events/hour | QC baseline; EHR baseline; Baseline + CPAP |
| T90% | Fraction of sleep-study time with $SpO_2$ below 90% | Sleep study | fraction | QC baseline; EHR baseline; Baseline + CPAP |
| T80% | Fraction of sleep-study time with $SpO_2$ below 80% | Sleep study | fraction | QC baseline; EHR baseline; Baseline + CPAP |
| AVERAGE_DESATURATION | Average oxygen saturation during desaturation episodes | Sleep study | % $SpO_2$ | QC baseline; EHR baseline; Baseline + CPAP |
| MEAN_SPO2 | Mean oxygen saturation | Sleep study | % $SpO_2$ | QC baseline; EHR baseline; Baseline + CPAP |
| MINIMUM_SPO2 | Minimum oxygen saturation | Sleep study | % $SpO_2$ | QC baseline; EHR baseline; Baseline + CPAP |
| MEAN_PULSE_RATE | Mean pulse rate during sleep study | Sleep study | beats/min | QC baseline; EHR baseline; Baseline + CPAP |
| MINIMUM_PULSE_RATE | Minimum pulse rate during sleep study | Sleep study | beats/min | QC baseline; EHR baseline; Baseline + CPAP |
| MAXIMUM_PULSE_RATE | Maximum pulse rate during sleep study | Sleep study | beats/min | QC baseline; EHR baseline; Baseline + CPAP |
| AHI_SUPINE | Supine apnea-hypopnea index | Sleep study | events/hour | QC baseline; EHR baseline; Baseline + CPAP |
| AHI_NONSUPINE | Non-supine apnea-hypopnea index | Sleep study | events/hour | QC baseline; EHR baseline; Baseline + CPAP |
| AHI | Overall apnea-hypopnea index using 4% criterion | Sleep study | events/hour | QC baseline; EHR baseline; Baseline + CPAP |
| **Questionnaire** | | | | |
| ESS1 | Epworth Sleepiness Scale item 1: Sitting and reading | Questionnaire | ordinal points | QC baseline; Baseline + CPAP |
| ESS2 | Epworth Sleepiness Scale item 2: Watching television | Questionnaire | ordinal points | QC baseline; Baseline + CPAP |
| ESS3 | Epworth Sleepiness Scale item 3: Sitting inactive in a public place | Questionnaire | ordinal points | QC baseline; Baseline + CPAP |
| ESS4 | Epworth Sleepiness Scale item 4: Passenger in a vehicle for one hour | Questionnaire | ordinal points | QC baseline; Baseline + CPAP |
| ESS5 | Epworth Sleepiness Scale item 5: Lying down to rest in the afternoon | Questionnaire | ordinal points | QC baseline; Baseline + CPAP |
| ESS6 | Epworth Sleepiness Scale item 6: Sitting and talking | Questionnaire | ordinal points | QC baseline; Baseline + CPAP |

| Raw feature | Plain-language definition | Source | Units/coding | Model membership* |
|---|---|---|---|---|
| ESS7 | Epworth Sleepiness Scale item 7: Sitting quietly after lunch | Questionnaire | ordinal points | QC baseline; Baseline + CPAP |
| ESS8 | Epworth Sleepiness Scale item 8: Stopped in traffic while driving | Questionnaire | ordinal points | QC baseline; Baseline + CPAP |
| GASP | Nocturnal gasping symptom | Questionnaire | ordinal points | QC baseline; Baseline + CPAP |
| SLEEPY | Daytime sleepiness symptom | Questionnaire | ordinal points | QC baseline; Baseline + CPAP |
| DROWSYDRIVING | Drowsy-driving symptom | Questionnaire | ordinal points | QC baseline; Baseline + CPAP |
| CLAUST | Claustrophobia symptom | Questionnaire | ordinal points | QC baseline; Baseline + CPAP |
| MEDS | Medication-related sleep item | Questionnaire | ordinal points | QC baseline; Baseline + CPAP |
| PARTNER | Bed-partner-related sleep item | Questionnaire | ordinal points | QC baseline; Baseline + CPAP |
| PARASOMNIA | Parasomnia symptom | Questionnaire | ordinal points | QC baseline; Baseline + CPAP |
| SLEEPATTACK | Sleep-attack symptom | Questionnaire | ordinal points | QC baseline; Baseline + CPAP |
| SLEEPHALLUCINATION | Sleep-related hallucination symptom | Questionnaire | ordinal points | QC baseline; Baseline + CPAP |
| CATAPLEXY | Cataplexy symptom indicator | Questionnaire | binary | QC baseline; Baseline + CPAP |
| ISI1 | Insomnia Severity Index item 1: Difficulty falling asleep | Questionnaire | ordinal points | QC baseline; Baseline + CPAP |
| ISI2 | Insomnia Severity Index item 2: Difficulty staying asleep | Questionnaire | ordinal points | QC baseline; Baseline + CPAP |
| ISI3 | Insomnia Severity Index item 3: Early awakening | Questionnaire | ordinal points | QC baseline; Baseline + CPAP |
| ISI4 | Insomnia Severity Index item 4: Satisfaction with sleep | Questionnaire | ordinal points | QC baseline; Baseline + CPAP |
| ISI5 | Insomnia Severity Index item 5: Interference with daily functioning | Questionnaire | ordinal points | QC baseline; Baseline + CPAP |
| ISI6 | Insomnia Severity Index item 6: Noticeability of sleep problem | Questionnaire | ordinal points | QC baseline; Baseline + CPAP |
| ISI7 | Insomnia Severity Index item 7: Distress or worry about sleep | Questionnaire | ordinal points | QC baseline; Baseline + CPAP |
| PHQ1 | PHQ-2 item 1: little interest or pleasure | Questionnaire | ordinal points | QC baseline; Baseline + CPAP |
| PHQ2 | PHQ-2 item 2: depressed mood | Questionnaire | ordinal points | QC baseline; Baseline + CPAP |
| FOSQ1 | Functional Outcomes of Sleep Questionnaire-10 item 1 | Questionnaire | raw ordinal points | QC baseline; Baseline + CPAP |
| FOSQ2 | Functional Outcomes of Sleep Questionnaire-10 item 2 | Questionnaire | raw ordinal points | QC baseline; Baseline + CPAP |
| FOSQ3 | Functional Outcomes of Sleep Questionnaire-10 item 3 | Questionnaire | raw ordinal points | QC baseline; Baseline + CPAP |
| FOSQ4 | Functional Outcomes of Sleep Questionnaire-10 item 4 | Questionnaire | raw ordinal points | QC baseline; Baseline + CPAP |
| FOSQ5 | Functional Outcomes of Sleep Questionnaire-10 item 5 | Questionnaire | raw ordinal points | QC baseline; Baseline + CPAP |
| FOSQ6 | Functional Outcomes of Sleep Questionnaire-10 item 6 | Questionnaire | raw ordinal points | QC baseline; Baseline + CPAP |
| FOSQ7 | Functional Outcomes of Sleep Questionnaire-10 item 7 | Questionnaire | raw ordinal points | QC baseline; Baseline + CPAP |
| FOSQ8 | Functional Outcomes of Sleep Questionnaire-10 item 8 | Questionnaire | raw ordinal points | QC baseline; Baseline + CPAP |
| FOSQ9 | Functional Outcomes of Sleep Questionnaire-10 item 9 | Questionnaire | raw ordinal points | QC baseline; Baseline + CPAP |
| FOSQ10 | Functional Outcomes of Sleep Questionnaire-10 item 10 | Questionnaire | raw ordinal points | QC baseline; Baseline + CPAP |
| PTSD | Post-traumatic-stress symptom indicator | Questionnaire | binary | QC baseline; Baseline + CPAP |
| SHIFT | Shift-work indicator | Questionnaire | binary | QC baseline; Baseline + CPAP |
| BEDTIME | Usual bedtime category | Questionnaire | nominal category; one-hot encoded | QC baseline; Baseline + CPAP |
| WAKETIME | Usual wake-time category | Questionnaire | nominal category; one-hot encoded | QC baseline; Baseline + CPAP |
| LATENCY | Self-reported sleep-onset latency | Questionnaire | minutes | QC baseline; Baseline + CPAP |
| RLS1 | Restless-legs symptom item 1 | Questionnaire | binary | QC baseline; Baseline + CPAP |
| RLS2 | Restless-legs symptom item 2 | Questionnaire | binary | QC baseline; Baseline + CPAP |
| RLS3 | Restless-legs symptom item 3 | Questionnaire | binary | QC baseline; Baseline + CPAP |
| SVS1 | Locally coded sleep-variable item 1 | Questionnaire | ordinal points | QC baseline; Baseline + CPAP |
| SVS2 | Locally coded sleep-variable item 2 | Questionnaire | ordinal points | QC baseline; Baseline + CPAP |
| NAPS | Napping item | Questionnaire | site-coded ordinal value | QC baseline; Baseline + CPAP |
| HEADACHE | Morning-headache item | Questionnaire | site-coded ordinal value | QC baseline; Baseline + CPAP |

| Raw feature | Plain-language definition | Source | Units/coding | Model membership* |
|---|---|---|---|---|
| AWAKENINGS | Nocturnal-awakenings item | Questionnaire | site-coded ordinal value | QC baseline; Baseline + CPAP |
| CAFFEINE | Caffeine-use item | Questionnaire | site-coded ordinal value | QC baseline; Baseline + CPAP |
| STOP1 | STOP item 1: snoring | Questionnaire | binary | QC baseline; Baseline + CPAP |
| STOP2 | STOP item 2: tiredness | Questionnaire | binary | QC baseline; Baseline + CPAP |
| STOP3 | STOP item 3: observed apnea | Questionnaire | binary | QC baseline; Baseline + CPAP |
| SLEEPTIME | Self-reported sleep duration | Questionnaire | hours/night | QC baseline; Baseline + CPAP |
| SLEEPPARALYSIS | Sleep-paralysis symptom frequency/severity | Questionnaire | ordinal points | QC baseline; Baseline + CPAP |
| CC_SNORING_APNEA | Snoring or observed-apnea chief-complaint flag | Questionnaire | binary | QC baseline; Baseline + CPAP |
| CC_INSOMNIA | Insomnia chief-complaint flag | Questionnaire | binary | QC baseline; Baseline + CPAP |
| CC_SLEEPINESS | Sleepiness chief-complaint flag | Questionnaire | binary | QC baseline; Baseline + CPAP |
| CC_FATIGUE | Fatigue chief-complaint flag | Questionnaire | binary | QC baseline; Baseline + CPAP |
| CC_RESTLESS_LEGS | Restless-legs chief-complaint flag | Questionnaire | binary | QC baseline; Baseline + CPAP |
| CC_PARASOMNIA | Parasomnia chief-complaint flag | Questionnaire | binary | QC baseline; Baseline + CPAP |
| CC_PAP_THERAPY | PAP-therapy chief-complaint flag | Questionnaire | binary | QC baseline; Baseline + CPAP |
| CC_HEADACHE | Headache chief-complaint flag | Questionnaire | binary | QC baseline; Baseline + CPAP |
| CC_DRY_MOUTH | Dry-mouth chief-complaint flag | Questionnaire | binary | QC baseline; Baseline + CPAP |
| CC_NOCTURIA | Nocturia chief-complaint flag | Questionnaire | binary | QC baseline; Baseline + CPAP |
| CC_REFERRAL | Referral-related chief-complaint flag | Questionnaire | binary | QC baseline; Baseline + CPAP |
| CC_OTHER | Other structured chief-complaint flag | Questionnaire | binary | QC baseline; Baseline + CPAP |
| ESS_comp | Epworth Sleepiness Scale total | Questionnaire | points | QC baseline; Baseline + CPAP |
| ISI_comp | Insomnia Severity Index total | Questionnaire | points | QC baseline; Baseline + CPAP |
| FOSQ_total | Functional Outcomes of Sleep Questionnaire-10 total | Questionnaire | points | QC baseline; Baseline + CPAP |
| general_productivity_score | FOSQ general-productivity domain score | Questionnaire | points | QC baseline; Baseline + CPAP |
| vigilance_score | FOSQ vigilance domain score | Questionnaire | points | QC baseline; Baseline + CPAP |
| social_outcome_score | FOSQ social-outcome domain score | Questionnaire | points | QC baseline; Baseline + CPAP |
| activity_level_score | FOSQ activity-level domain score | Questionnaire | points | QC baseline; Baseline + CPAP |
| intimacy_score | FOSQ intimacy/sexual-relationships domain score | Questionnaire | points | QC baseline; Baseline + CPAP |
| **CPAP** | | | | |
| mean_hrs_night | Mean PAP use across every calendar night in the early window | CPAP | hours/night | Baseline + CPAP; CPAP only |
| pct_nights_used | Percentage of early-window nights with PAP use >0 | CPAP | percentage points | Baseline + CPAP; CPAP only |
| pct_nights_ge4h | Percentage of early-window nights with PAP use ≥4 hours | CPAP | percentage points | Baseline + CPAP; CPAP only |
| mean_usage_used | Mean PAP use on used nights | CPAP | minutes/used night | Baseline + CPAP; CPAP only |
| mean_leak95_lpm | Mean nightly 95th-percentile leak over used nights | CPAP | L/min | Baseline + CPAP; CPAP only |
| mean_p95_cmh2o | Mean nightly 95th-percentile pressure over used nights | CPAP | $cmH_2O$ | Baseline + CPAP; CPAP only |
| mean_rahi | Mean nightly residual apnea-hypopnea index over used nights | CPAP | events/hour | Baseline + CPAP; CPAP only |

**Supplement S3 Phase 2 sensitivity analysis comparing setup-date- and first-use-anchored cumulative CPAP-data windows for the primary Month-12 outcome, mean use ≥4 hours/night.**

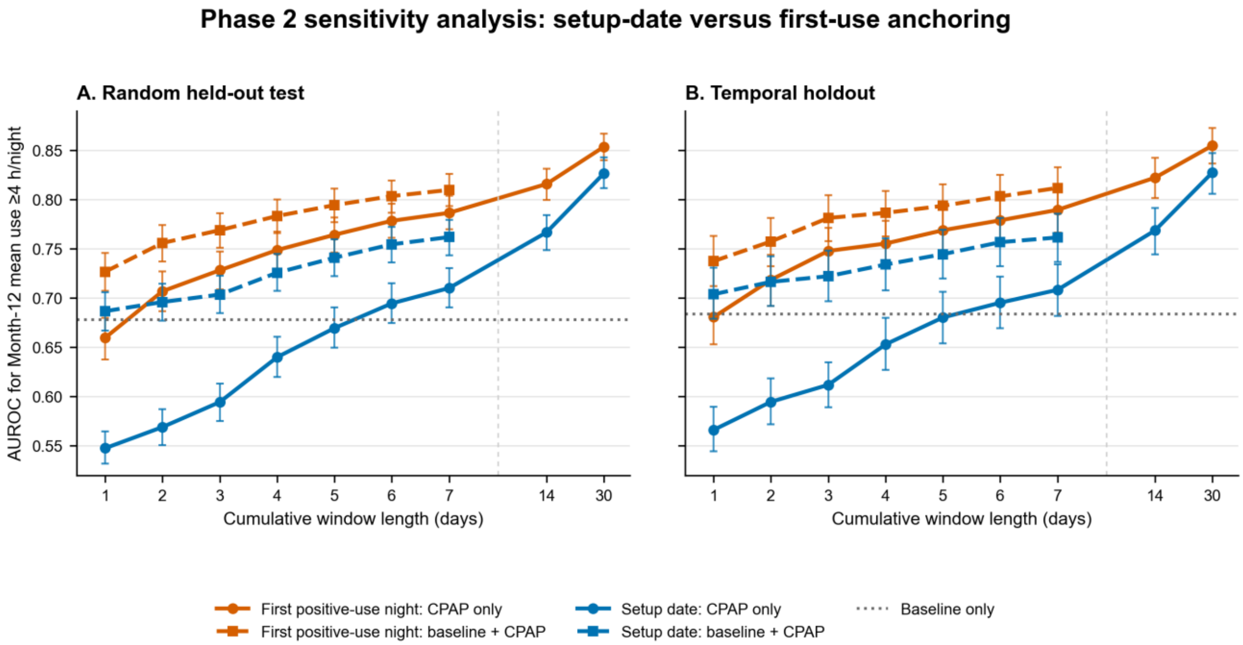


Supplement S3. Phase 2 sensitivity analysis comparing setup-date- and first-use-anchored cumulative CPAP-data windows for the primary Month-12 outcome, mean use ≥4 hours/night. Panels show (A) the random held-out test sample (n=2,982) and (B) the temporal holdout (n=1,676). Dotted horizontal lines show baseline-only XGBoost; solid lines show CPAP-only models and dashed lines show baseline+CPAP models. Baseline+CPAP models were evaluated through day 7; day-14 and day-30 points are CPAP-only. Error bars are bootstrap 95% confidence intervals. AUROC indicates area under the receiver-operating-characteristic curve.

### Supplement S4. Phase 4 reduced-feature model definitions

- **Parsimonious baseline model:** 13 raw variables—AHI, age, sleep-study analysis duration, race/ethnicity, Elixhauser comorbidity index, weight, height, depression, ODI4%, influenza vaccination, sex, mean pulse rate, and BMI. Race/ethnicity was expanded into indicator variables, yielding 21 encoded predictors. Variables were selected based on data availability, importance in the full KPSC models, and clinician judgment.
- **Questionnaire–Sleep-Study baseline model:** 98 raw variables comprising 76 questionnaire variables, 18 diagnostic sleep-study variables, and age, sex, height, and weight; these yielded 120 encoded predictors. This model excluded other EHR-derived clinical, utilization, medication, and comorbidity variables.
- **Early CPAP augmentation:** Each reduced baseline model was evaluated alone and after addition of the same eight CPAP-data summaries from days 1–7 used in Phases 2 and 3.
- **CPAP-data-only comparator:** The common CPAP-data-only model was used across analyses and was not refitted separately for each baseline feature family.

### Supplement S5 Phase 1 univariate demographic associations with the primary outcome, Month-12 mean CPAP use ≥4 hours/night

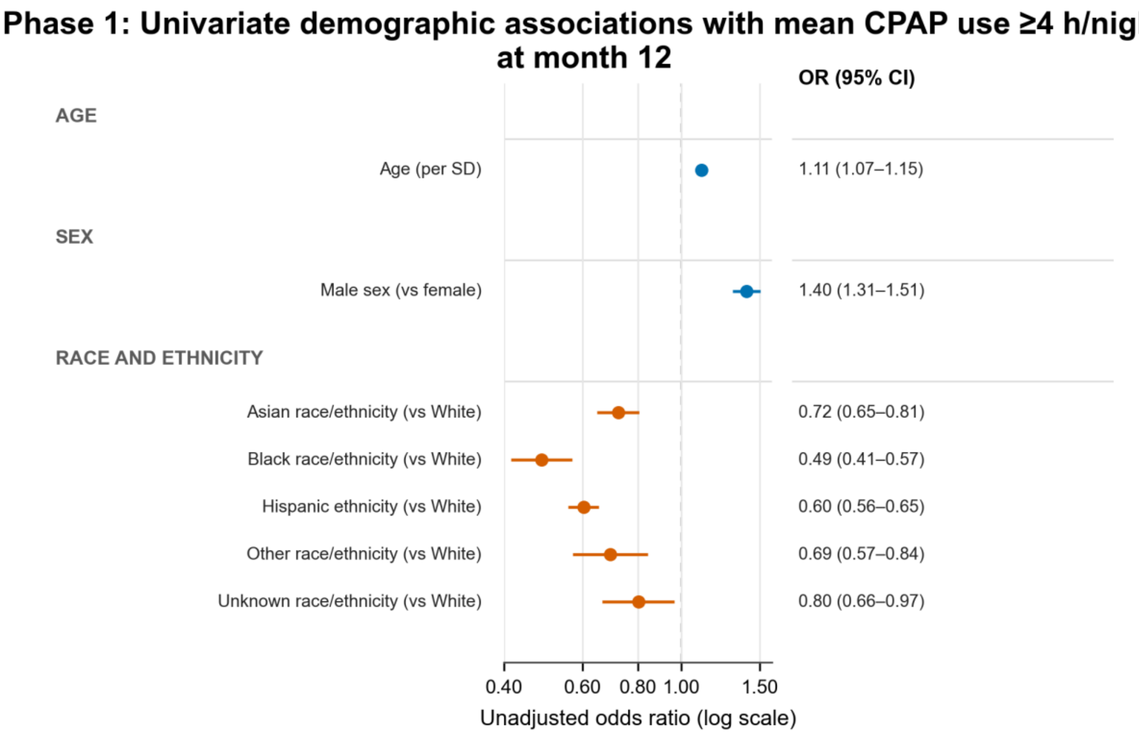


Supplement S5. Phase 1 univariate demographic associations with the primary outcome, Month-12 mean CPAP use ≥4 hours/night, in the Questionnaire-Complete earlier-date cohort (n=14,906). Unadjusted odds ratios and 95% confidence intervals are from separate logistic-regression models for age, sex, and race and ethnicity. Age is scaled per SD (13.85 years); female sex and White race/ethnicity are reference categories. The race-and-ethnicity omnibus likelihood-ratio test was P<.001.

### Supplement S6 Phase 1 pre-initiation factors associated with CMS 90-day adherence.

**A**

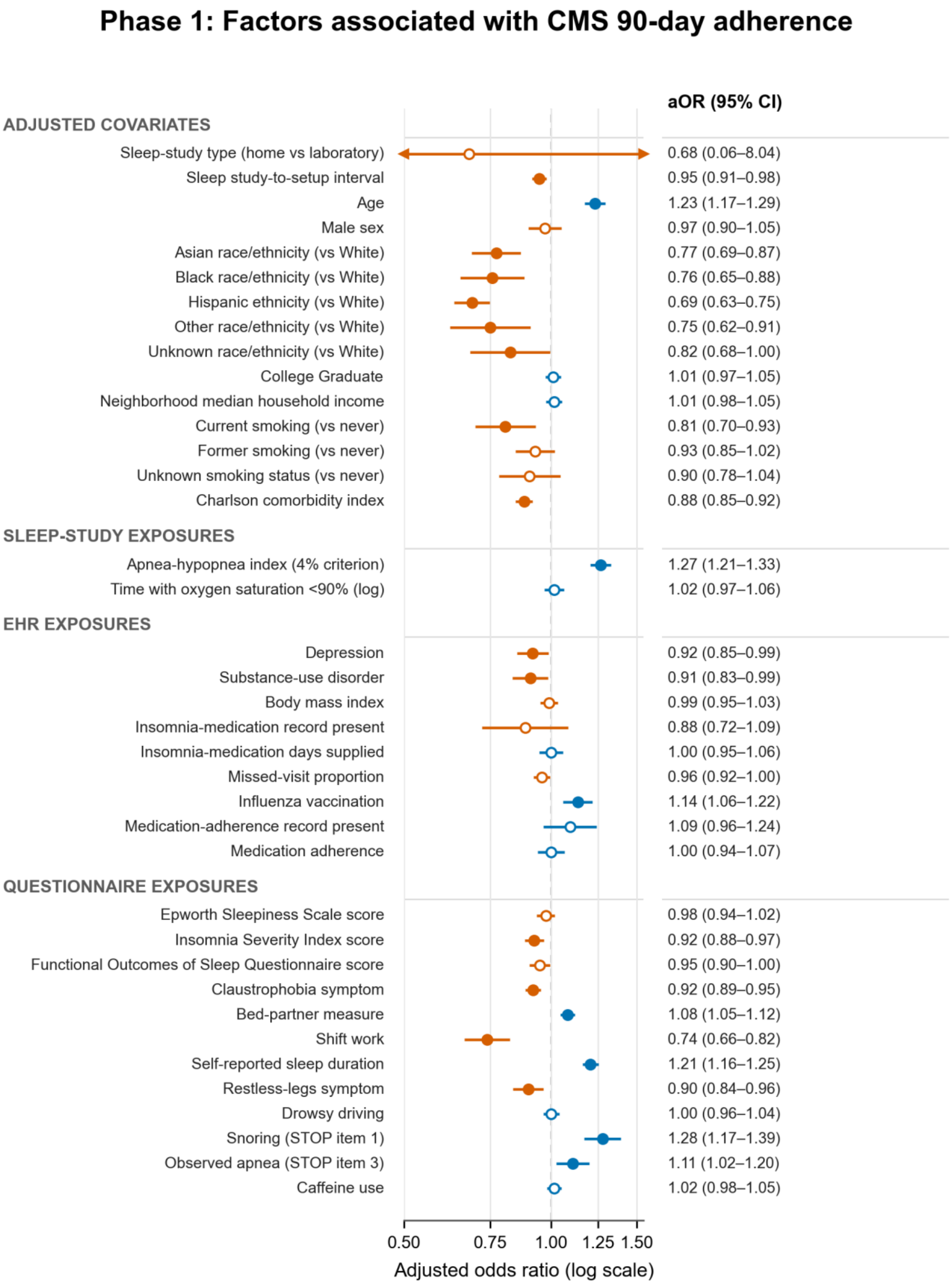

**B**

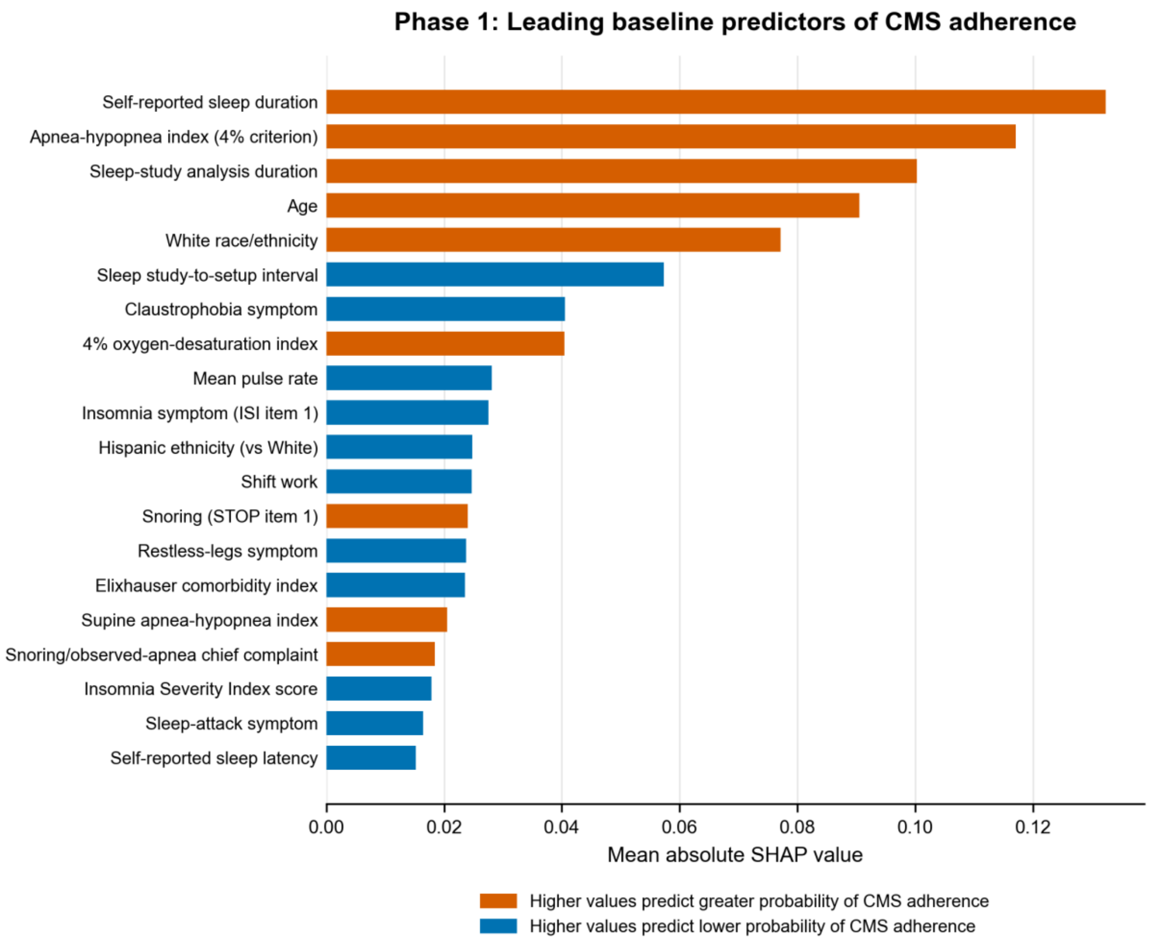


Supplement S6. Phase 1 pre-initiation factors associated with CMS 90-day adherence. (A) Adjusted odds ratios and 95% confidence intervals from the prespecified logistic-regression model. (B) Mean absolute SHAP values for the leading baseline XGBoost features.

**Supplement S7 Phase 1 pre-initiation factors associated with Month-12 mean CPAP use ≥2 hours/night.**

**A**

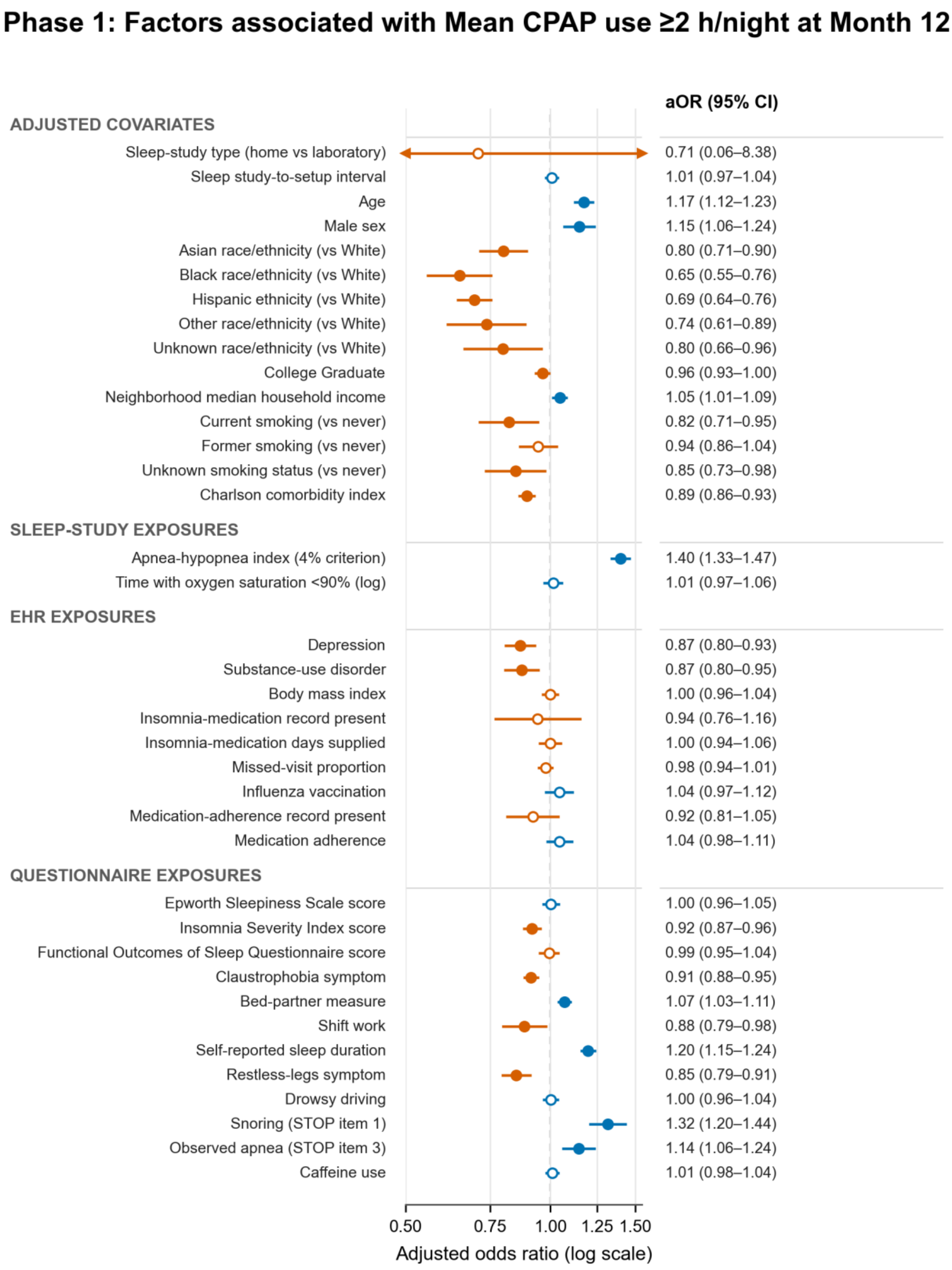

B

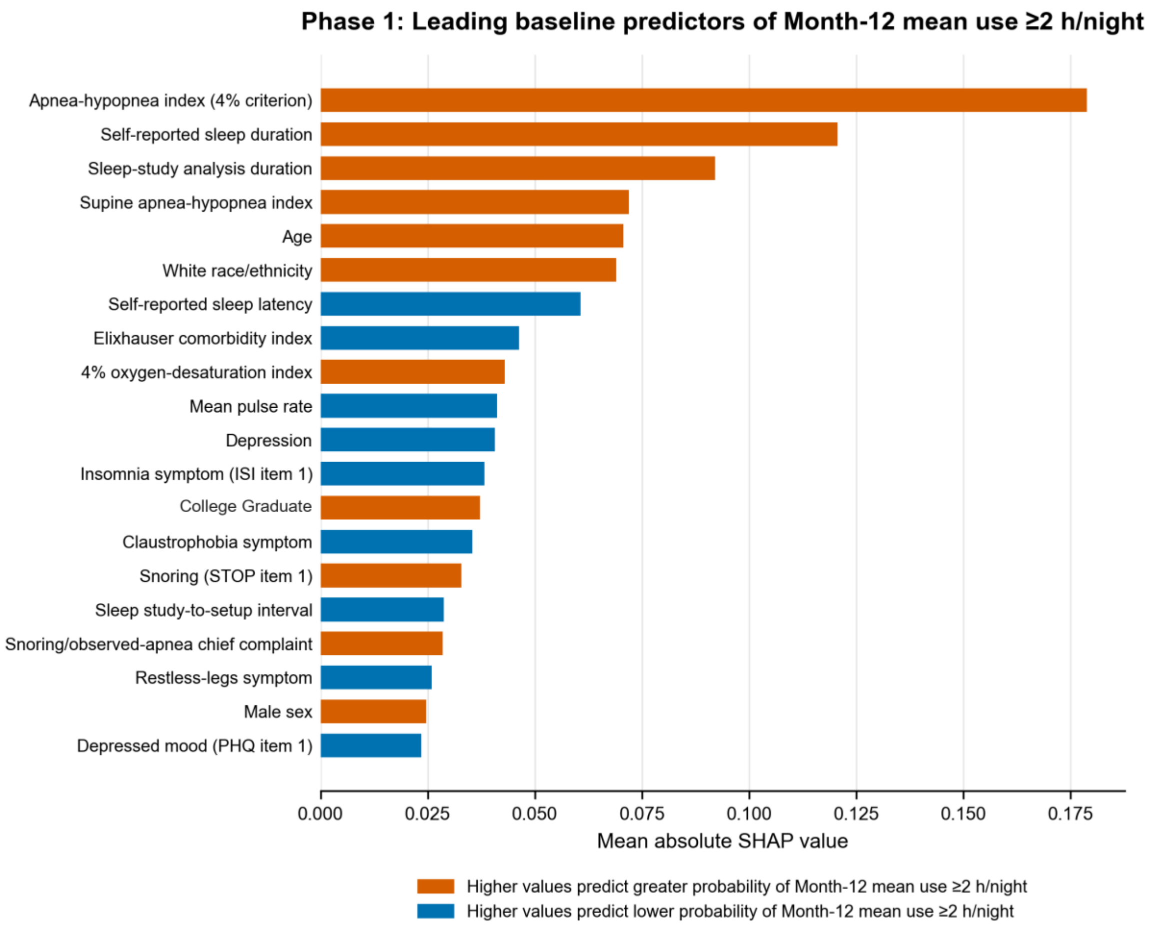


Supplement S7. Phase 1 pre-initiation factors associated with Month-12 mean CPAP use ≥2 hours/night. (A) Adjusted odds ratios and 95% confidence intervals from the prespecified logistic-regression model. (B) Mean absolute SHAP values for the leading baseline XGBoost features.

**Supplement S8 Phase 1 pre-initiation factors associated with Month-12 treatment abandonment**

**A**

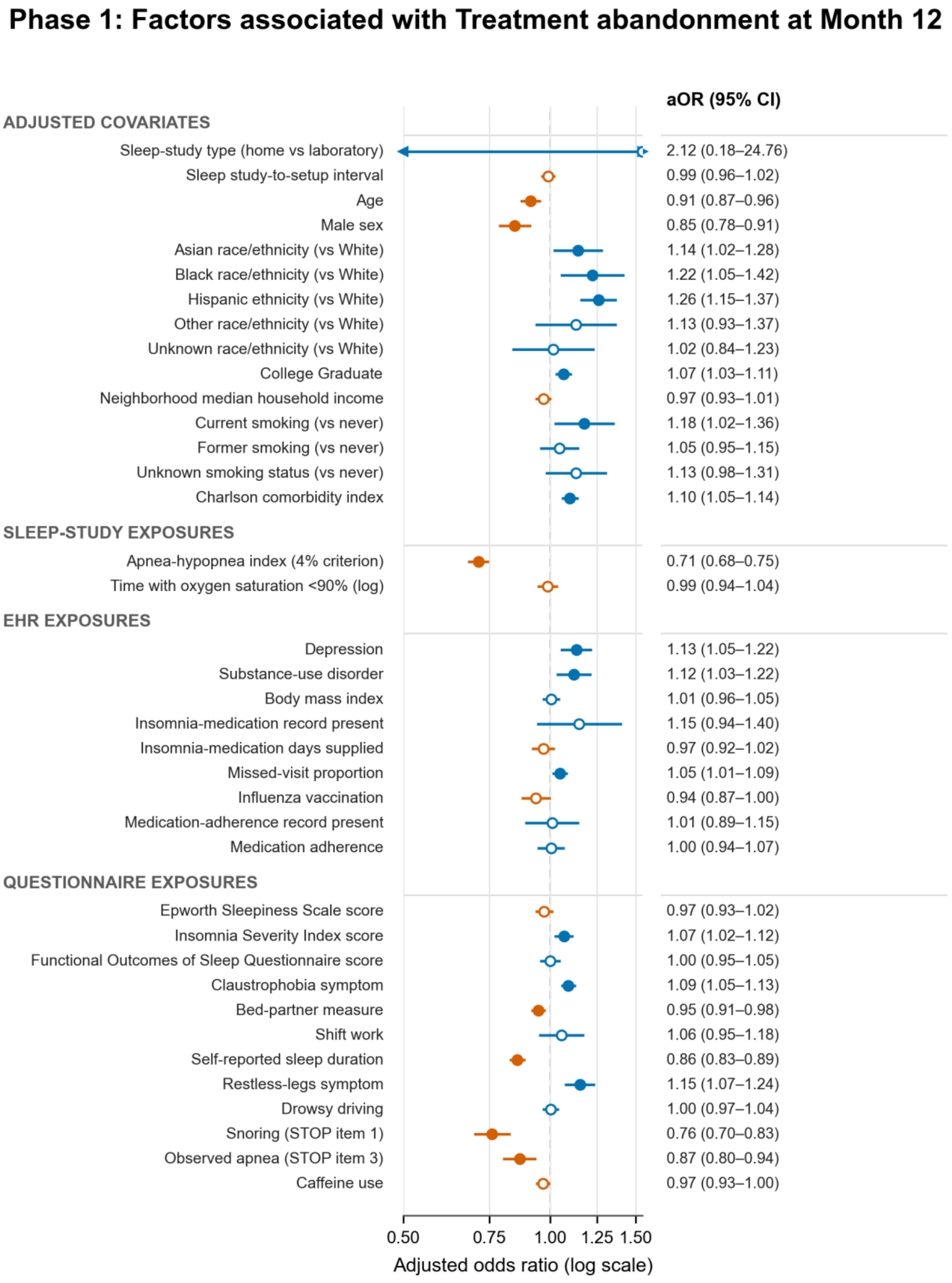

**B**

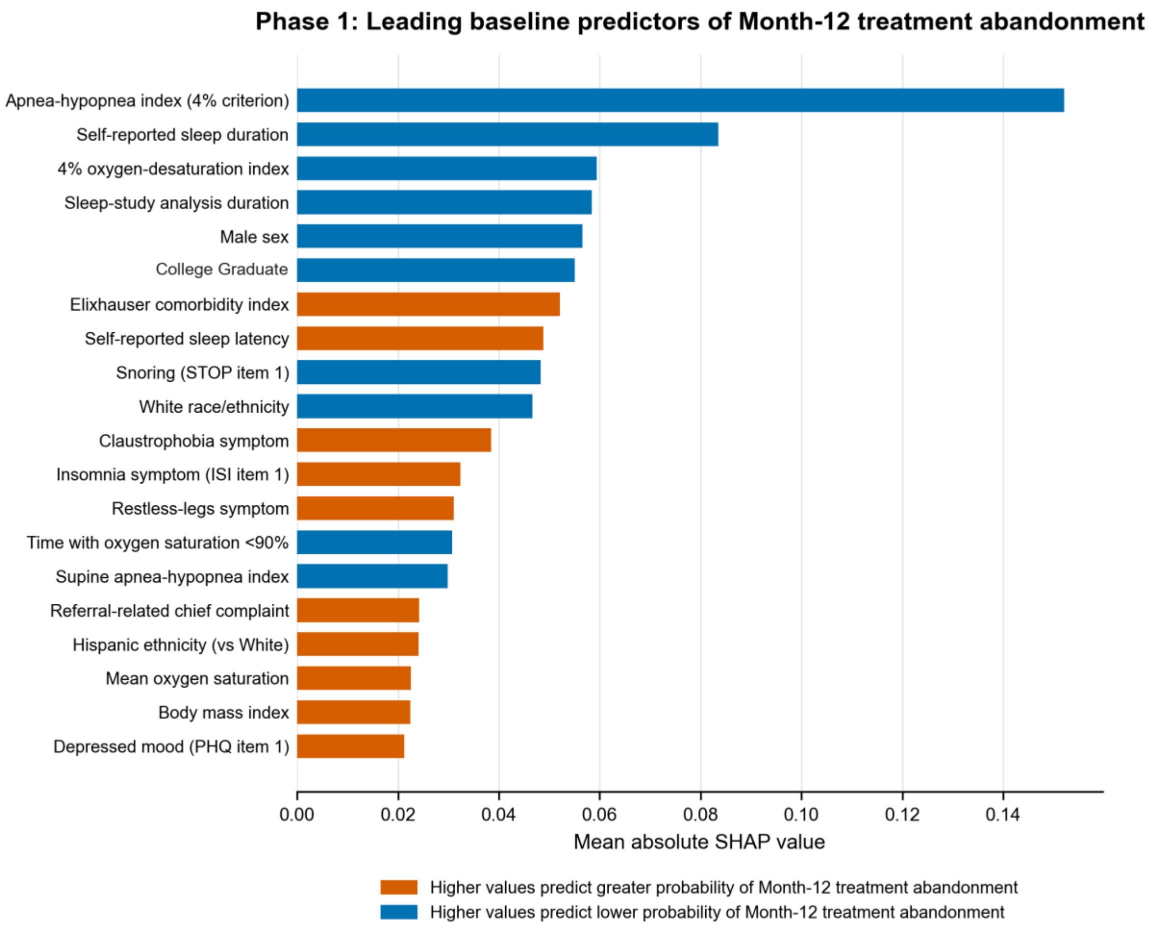


Supplement S8. Phase 1 pre-initiation factors associated with Month-12 treatment abandonment, defined as no recorded use during days 331–360. (A) Adjusted odds ratios and 95% confidence intervals, reoriented so values greater than 1 indicate greater odds of abandonment. (B) Mean absolute SHAP values from the any-use XGBoost model, with direction reoriented to abandonment.

**Supplement S9 Temporal calibration of Phase 2 XGBoost models for the primary Month-12 outcome, mean CPAP use ≥4 hours/night**

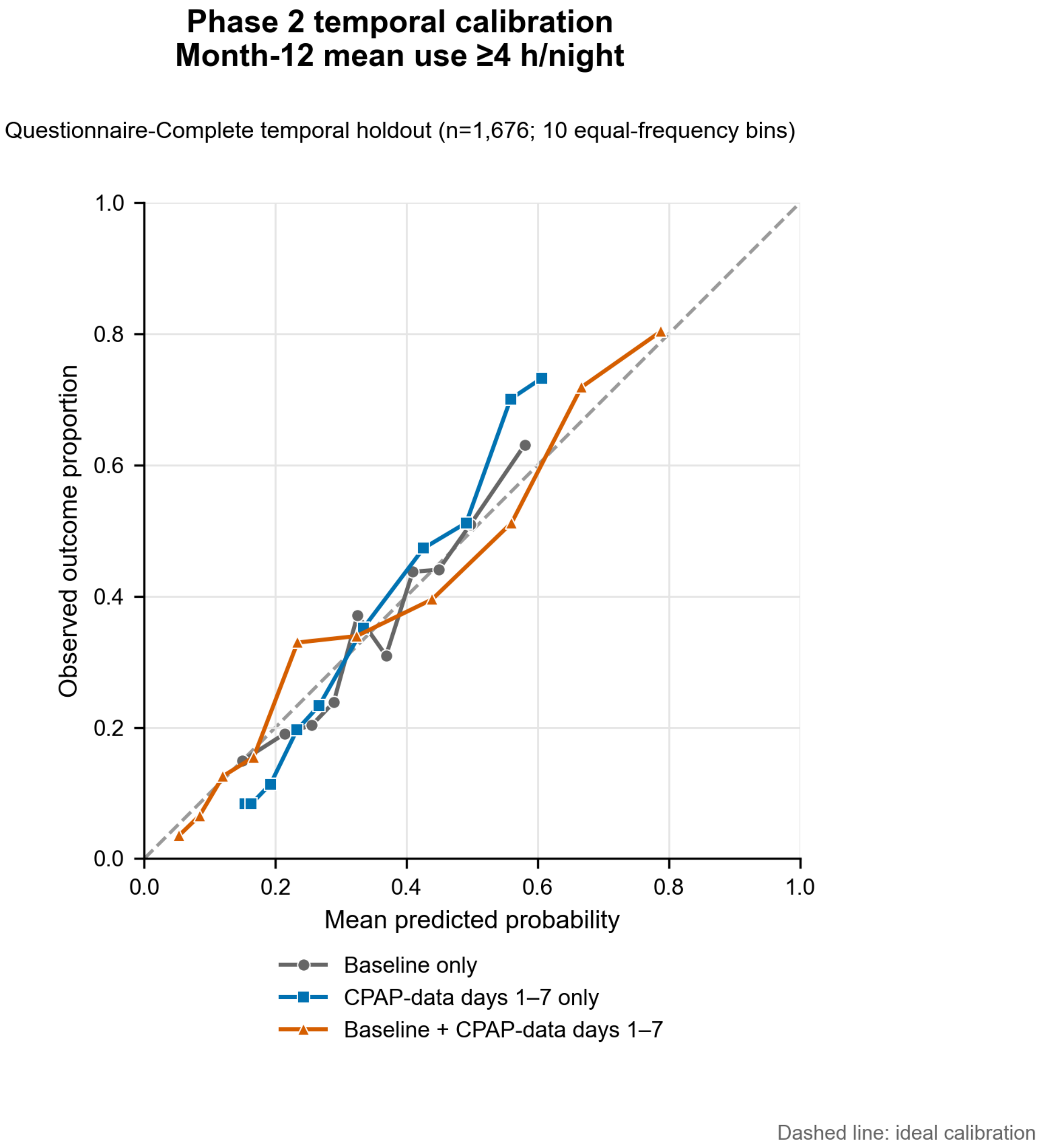


Supplement S9. Temporal calibration of Phase 2 XGBoost models for the primary Month-12 outcome, mean CPAP use ≥4 hours/night, in the Questionnaire-Complete temporal holdout (n=1,676). Points show observed outcome frequency versus mean predicted probability within 10 equal-frequency groups for baseline-only, CPAP-data days 1–7 only, and baseline+CPAP-data days 1–7 models; lines connect the groups. The diagonal line indicates ideal calibration. Model probabilities were not recalibrated in the temporal holdout.

### Supplement S10 Temporal one-versus-rest calibration of Phase 3 XGBoost models for day-8-to-364 CPAP trajectory class

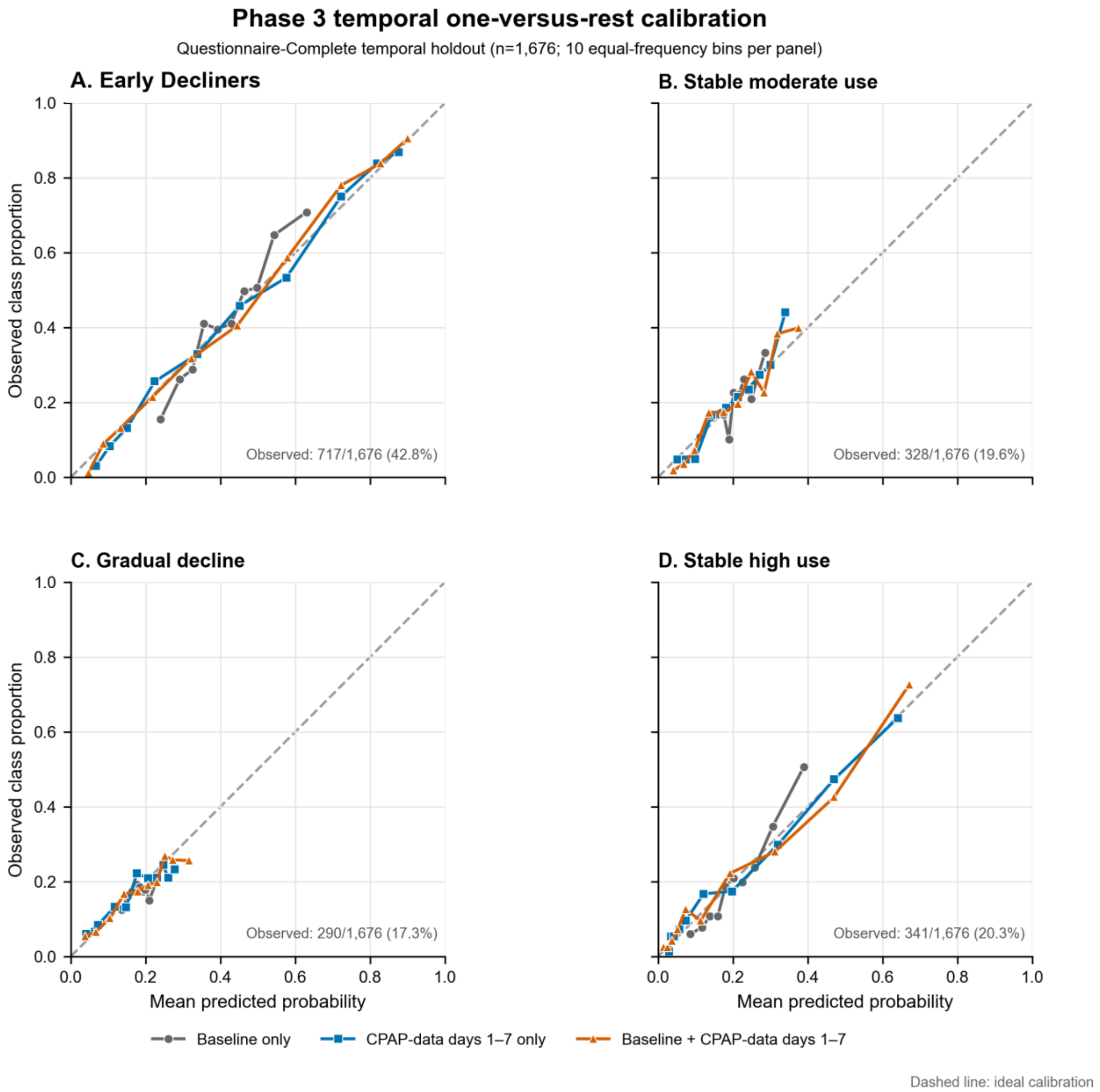


Supplement S10. Temporal one-versus-rest calibration of Phase 3 XGBoost models for day-8-to-364 CPAP trajectory class in the Questionnaire-Complete temporal holdout (n=1,676). Panels show (A) Early Decliners, (B) stable moderate use, (C) gradual decline, and (D) stable high use. Points show observed class frequency versus mean predicted probability within 10 equal-frequency groups for baseline-only, CPAP-data days 1–7 only, and baseline+ CPAP-data days 1–7 models; lines connect the groups. The diagonal line indicates ideal calibration. Model probabilities were not recalibrated in the temporal holdout.

### Supplement S11. One-versus-rest AUROC for prediction of day-8-to-364 CPAP trajectory class.

| Model | Macro average | Early Decliners | Stable moderate | Gradual decline | Stable high |
|---|---|---|---|---|---|
| **Random held-out test (n=2,981)** | | | | | |
| Baseline | 0.619 (0.605–0.632) | 0.667 (0.647–0.685) | 0.584 (0.559–0.610) | 0.533 (0.506–0.561) | 0.691 (0.667–0.713) |
| CPAP days 1–7 only | 0.756 (0.744–0.767) | 0.861 (0.848–0.873) | 0.673 (0.649–0.696) | 0.646 (0.621–0.669) | 0.843 (0.826–0.860) |
| Baseline + CPAP | 0.770 (0.759–0.781) | 0.871 (0.859–0.884) | 0.691 (0.668–0.714) | 0.657 (0.633–0.681) | 0.859 (0.843–0.874) |
| **Temporal holdout (n=1,676)** | | | | | |
| Baseline | 0.632 (0.614–0.649) | 0.686 (0.663–0.711) | 0.604 (0.570–0.637) | 0.524 (0.488–0.560) | 0.715 (0.686–0.744) |
| CPAP days 1–7 only | 0.748 (0.732–0.763) | 0.849 (0.831–0.867) | 0.710 (0.680–0.739) | 0.614 (0.581–0.646) | 0.818 (0.794–0.842) |
| Baseline + CPAP | 0.767 (0.751–0.782) | 0.864 (0.847–0.881) | 0.725 (0.697–0.754) | 0.645 (0.610–0.676) | 0.833 (0.810–0.856) |

Values are AUROC (95% CI). The four class-specific columns are one-versus-rest estimates; macro average is the unweighted mean across classes. CPAP predictors summarize calendar days 1–7 beginning with the first positive-use night. AUROC indicates area under the receiver-operating-characteristic curve; CI, confidence interval.

### Supplement S12. Temporal calibration measures for Phase 2 primary-outcome and Phase 3 trajectory-class prediction.

| Target / class | Model | N | Observed, n (%) | Calibration-in-the-large (95% CI) | Calibration slope (95% CI) | Brier score |
|---|---|---|---|---|---|---|
| **Phase 2—Primary Month-12 outcome (mean use ≥4 hours/night)** | | | | | | |
| Primary outcome | Baseline only | 1,676 | 583 (34.8%) | −0.03 (−0.13–0.08) | 1.17 (0.98–1.37) | 0.206 |
| Primary outcome | CPAP-data days 1–7 only | 1,676 | 583 (34.8%) | 0.03 (−0.08–0.13) | 1.56 (1.40–1.73) | 0.178 |
| Primary outcome | Baseline + CPAP-data days 1–7 | 1,676 | 583 (34.8%) | 0.03 (−0.09–0.14) | 1.01 (0.90–1.11) | 0.165 |
| **Phase 3—Day-8-to-364 trajectory class (one versus rest)** | | | | | | |
| Early Decliners | Baseline only | 1,676 | 717 (42.8%) | 0.05 (−0.05–0.15) | 1.45 (1.22–1.67) | 0.221 |
| Early Decliners | CPAP-data days 1–7 only | 1,676 | 717 (42.8%) | −0.03 (−0.15–0.09) | 1.07 (0.97–1.17) | 0.156 |
| Early Decliners | Baseline + CPAP-data days 1–7 | 1,676 | 717 (42.8%) | −0.00 (−0.12–0.12) | 1.08 (0.98–1.18) | 0.147 |
| Stable moderate | Baseline only | 1,676 | 328 (19.6%) | −0.00 (−0.12–0.12) | 1.18 (0.78–1.57) | 0.154 |
| Stable moderate | CPAP-data days 1–7 only | 1,676 | 328 (19.6%) | 0.03 (−0.10–0.15) | 1.27 (1.04–1.50) | 0.144 |
| Stable moderate | Baseline + CPAP-data days 1–7 | 1,676 | 328 (19.6%) | 0.00 (−0.12–0.13) | 1.22 (1.01–1.43) | 0.143 |
| Gradual decline | Baseline only | 1,676 | 290 (17.3%) | −0.06 (−0.18–0.07) | 0.52 (−0.16–1.20) | 0.143 |
| Gradual decline | CPAP-data days 1–7 only | 1,676 | 290 (17.3%) | −0.03 (−0.16–0.10) | 0.74 (0.51–0.97) | 0.140 |
| Gradual decline | Baseline + CPAP-data days 1–7 | 1,676 | 290 (17.3%) | −0.05 (−0.18–0.08) | 0.81 (0.60–1.03) | 0.138 |
| Stable high | Baseline only | 1,676 | 341 (20.3%) | −0.02 (−0.14–0.10) | 1.50 (1.25–1.75) | 0.147 |
| Stable high | CPAP-data days 1–7 only | 1,676 | 341 (20.3%) | 0.04 (−0.10–0.18) | 0.93 (0.83–1.04) | 0.123 |
| Stable high | Baseline + CPAP-data days 1–7 | 1,676 | 341 (20.3%) | 0.06 (−0.08–0.21) | 0.94 (0.83–1.05) | 0.117 |

Calibration-in-the-large was estimated with the predicted log-odds as an offset (ideal, 0); calibration slope was estimated from a logistic recalibration model containing an intercept and predicted log-odds (ideal, 1). Parenthetical values are 95% Wald confidence intervals. Brier scores were calculated from the original, unrecalibrated probabilities; lower values indicate better probabilistic accuracy. Phase 3 Brier scores are one versus rest; corresponding multiclass Brier scores for baseline-only, CPAP-data days 1–7 only, and combined models were 0.666, 0.564, and 0.545, respectively. Calibration curves used 10 equal-frequency groups. CPAP-data days 1–7 began with the first positive-use night. CI indicates confidence interval.

### Supplement S13. Baseline-only logistic-regression and XGBoost AUROCs for the four Phase 2 outcomes.

| Phase 2 outcome | Logistic regression AUROC (95% CI) | XGBoost AUROC (95% CI) |
|---|---|---|
| Month-12 mean use ≥4 hours/night | 0.672 (0.652–0.692) | 0.678 (0.657–0.697) |
| Month-12 mean use ≥2 hours/night | 0.668 (0.648–0.687) | 0.676 (0.656–0.694) |
| Any Month-12 use | 0.643 (0.623–0.663) | 0.658 (0.639–0.678) |
| CMS 90-day adherence | 0.657 (0.637–0.676) | 0.653 (0.633–0.673) |

Values are AUROC (95% CI) from baseline-only models in the Questionnaire-Complete KPSC random held-out test set (n=2,982). Both model families used pretreatment EHR, sleep-study, and questionnaire data; no CPAP-use data were included. Confidence intervals were calculated using 2,000 stratified bootstrap resamples. Month-12 outcomes refer to days 331–360; CMS 90-day adherence requires ≥4 hours/night on ≥70% of nights during a consecutive 30-day interval within the first 90 days after setup. AUROC indicates area under the receiver-operating-characteristic curve; CI, confidence interval; CMS, Centers for Medicare & Medicaid Services.

### Supplement S14 Baseline decision-curve analysis

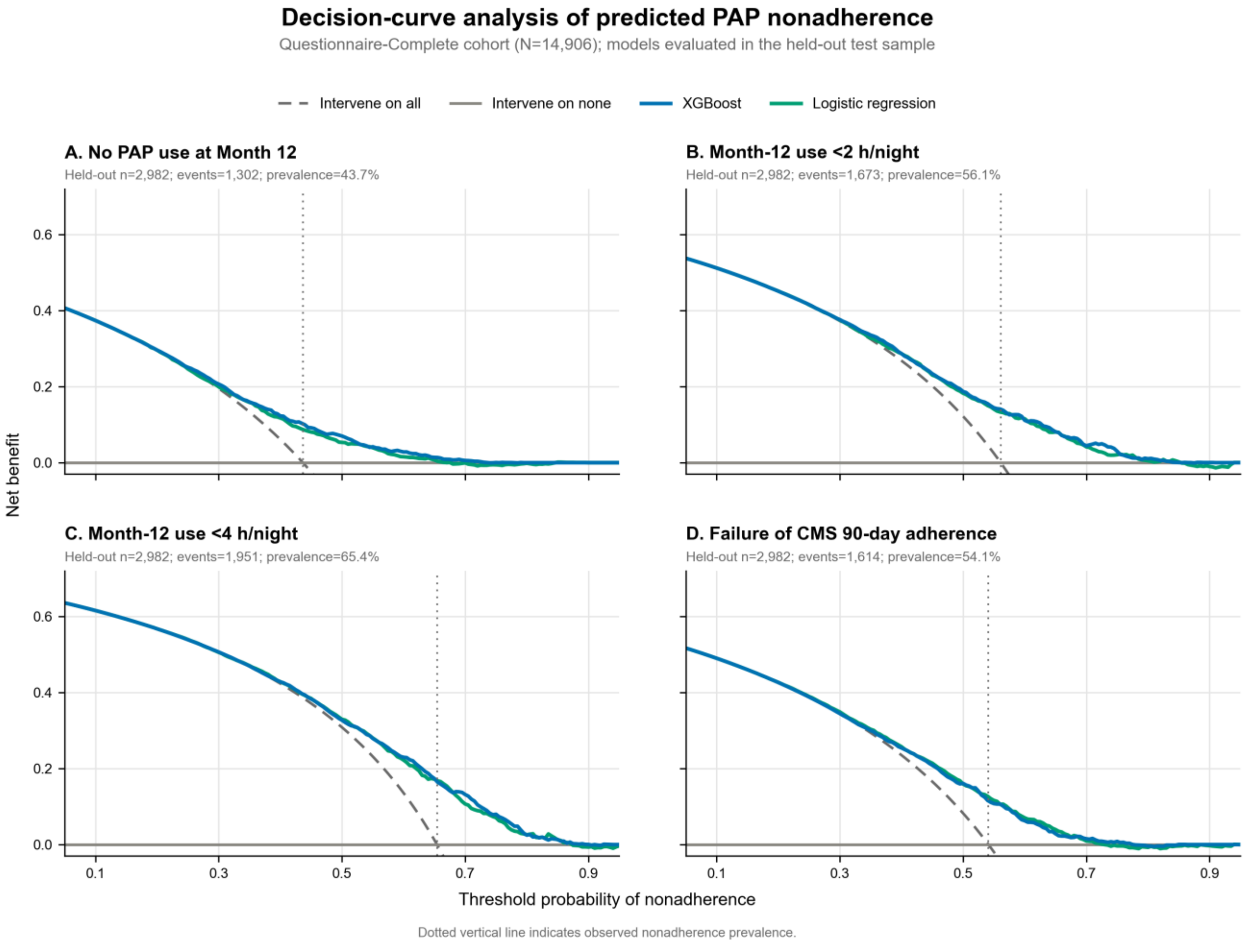


Supplement S14. Baseline decision-curve analysis for predicted nonadherence in the Questionnaire-Complete random held-out test sample (n=2,982). Panels show (A) no PAP use at Month 12, (B) Month-12 mean use <2 hours/night, (C) Month-12 mean use <4 hours/night, and (D) failure of CMS 90-day adherence. Patients were assigned to an intervention when predicted nonadherence risk equaled or exceeded the threshold probability. Curves compare XGBoost and logistic-regression net benefit with intervene-all and intervene-none strategies.

### Supplement S15. Baseline and first-90-day CPAP characteristics by day-8-to-364 CPAP-use trajectory class, with four-group omnibus comparisons.

| Characteristic | Overall (N=16,582) | Early Decliners (N=7,116) | Stable moderate use (N=3,147) | Gradual decline (N=2,888) | Stable high use (N=3,431) | P value* |
|---|---|---|---|---|---|---|
| N | 16,582 | 7,116 | 3,147 | 2,888 | 3,431 | |
| **Demographics** | | | | | | |
| Age, years | 49.6 ± 13.8 | 48.3 ± 13.8 | 49.8 ± 13.3 | 49.9 ± 13.5 | 51.9 ± 14.3 | <0.001 |
| Sex: Female | 6,078 (36.7%) | 2,788 (39.2%) | 1,030 (32.7%) | 1,122 (38.9%) | 1,138 (33.2%) | <0.001 |
| Sex: Male | 10,504 (63.3%) | 4,328 (60.8%) | 2,117 (67.3%) | 1,766 (61.1%) | 2,293 (66.8%) | |
| Sex: Other/unknown | 0 (0.0%) | 0 (0.0%) | 0 (0.0%) | 0 (0.0%) | 0 (0.0%) | |
| Race/ethnicity: Asian | 2,082 (12.6%) | 886 (12.5%) | 455 (14.5%) | 398 (13.8%) | 343 (10.0%) | <0.001 |
| Race/ethnicity: Black | 1,021 (6.2%) | 515 (7.2%) | 190 (6.0%) | 184 (6.4%) | 132 (3.8%) | |
| Race/ethnicity: Hispanic | 6,174 (37.2%) | 2,991 (42.0%) | 1,107 (35.2%) | 1,075 (37.2%) | 1,001 (29.2%) | |
| Race/ethnicity: White | 6,137 (37.0%) | 2,234 (31.4%) | 1,174 (37.3%) | 1,017 (35.2%) | 1,712 (49.9%) | |
| Race/ethnicity: Other/Multiple/Unknown | 1,168 (7.0%) | 490 (6.9%) | 221 (7.0%) | 214 (7.4%) | 243 (7.1%) | |
| **Socioeconomic context** | | | | | | |
| Area-level median household income: <= $45,000 | 455 (2.7%) | 227 (3.2%) | 68 (2.2%) | 90 (3.1%) | 70 (2.0%) | <0.001 |
| Area-level median household income: $45,001-$80,000 | 4,319 (26.0%) | 2,020 (28.4%) | 744 (23.6%) | 767 (26.6%) | 788 (23.0%) | |
| Area-level median household income: $80,001+ | 7,626 (46.0%) | 3,099 (43.5%) | 1,543 (49.0%) | 1,278 (44.3%) | 1,706 (49.7%) | |
| Area-level median household income: Missing/unknown | 4,182 (25.2%) | 1,770 (24.9%) | 792 (25.2%) | 753 (26.1%) | 867 (25.3%) | |
| College Graduate | 20.2 ± 10.2 [4,188 missing] | 19.3 ± 10.2 [1,771 missing] | 21.0 ± 10.2 [795 missing] | 20.0 ± 10.2 [755 missing] | 21.5 ± 10.2 [867 missing] | <0.001 |
| **Clinical and questionnaire** | | | | | | |
| AHI, events/h | 34.2 ± 26.0 | 30.2 ± 25.1 | 37.7 ± 26.4 | 33.6 ± 24.6 | 40.0 ± 27.4 | <0.001 |
| BMI, kg/$m^2$ | 35.8 ± 8.4 [3,241 missing] | 36.0 ± 8.5 [1,283 missing] | 35.6 ± 8.2 [639 missing] | 35.4 ± 8.1 [587 missing] | 35.9 ± 8.3 [732 missing] | 0.022 |
| Charlson index score | 1.3 ± 1.5 | 1.4 ± 1.5 | 1.3 ± 1.4 | 1.4 ± 1.5 | 1.3 ± 1.4 | <0.001 |
| Depression | 7,298 (44.0%) | 3,444 (48.4%) | 1,207 (38.4%) | 1,297 (44.9%) | 1,350 (39.3%) | <0.001 |
| Epworth Sleepiness Scale score (0–24) | 9.2 ± 5.2 | 9.3 ± 5.3 | 9.5 ± 5.1 | 9.3 ± 5.3 | 8.6 ± 5.0 | <0.001 |
| Insomnia Severity Index score (0–28) | 14.0 ± 5.4 | 14.7 ± 5.4 | 13.5 ± 5.2 | 14.0 ± 5.3 | 13.0 ± 5.3 | <0.001 |
| Self-reported sleep duration, hours/night | 6.1 ± 1.3 | 5.9 ± 1.3 | 6.1 ± 1.2 | 6.1 ± 1.2 | 6.5 ± 1.2 | <0.001 |
| **90-day CPAP adherence (descriptive)** | | | | | | |
| CMS Adherence Rate | 7,501 (45.2%) | 561 (7.9%) | 2,164 (68.8%) | 1,711 (59.2%) | 3,065 (89.3%) | <0.001 |
| Mean hours/use (first 90 days) | 3.5 ± 2.6 | 1.2 ± 1.2 | 4.7 ± 1.6 | 4.1 ± 1.5 | 6.8 ± 1.3 | <0.001 |
| % used 4+ hours (first 90 days) | 41.8 ± 34.7 | 9.9 ± 12.7 | 60.4 ± 24.0 | 50.2 ± 21.7 | 83.6 ± 18.0 | <0.001 |

Values are mean ± SD or n (%). The table pools the Questionnaire-Complete earlier-date and temporal cohorts (N=16,582) and uses mutually exclusive trajectory classes assigned by the frozen four-class model. Percentages use the full column denominator; bracketed values report missing observations. *P values are unadjusted omnibus comparisons from one-way analysis of variance for continuous variables and Pearson chi-square tests for categorical variables; missing continuous observations were excluded. For multilevel categorical variables, the omnibus P value is shown on the first listed level. AHI indicates apnea–hypopnea index; BMI, body mass index; CMS, Centers for Medicare & Medicaid Services.

### Supplement S16. Conditional pairwise AUROC for prediction of day-8-to-364 CPAP trajectory class.

| Class A | Class B | N | Baseline | CPAP days 1-7 only | Baseline + CPAP |
|---|---|---|---|---|---|
| **Random held-out test (n=2,981)** | | | | | |
| Early Decliners | Stable moderate | 1,840 | 0.656 (0.630-0.683) | 0.829 (0.809-0.849) | 0.845 (0.824-0.864) |
| Early Decliners | Gradual decline | 1,778 | 0.592 (0.562-0.620) | 0.811 (0.789-0.832) | 0.817 (0.796-0.838) |
| Early Decliners | Stable high | 1,913 | 0.740 (0.716-0.763) | 0.929 (0.915-0.942) | 0.942 (0.931-0.952) |
| Stable moderate | Gradual decline | 1,068 | 0.571 (0.537-0.605) | 0.543 (0.510-0.576) | 0.581 (0.546-0.615) |
| Stable moderate | Stable high | 1,203 | 0.620 (0.589-0.650) | 0.740 (0.711-0.767) | 0.756 (0.728-0.783) |
| Gradual decline | Stable high | 1,141 | 0.656 (0.625-0.688) | 0.755 (0.727-0.783) | 0.779 (0.752-0.806) |
| **Temporal holdout (n=1,676)** | | | | | |
| Early Decliners | Stable moderate | 1,045 | 0.680 (0.646-0.713) | 0.838 (0.814-0.863) | 0.858 (0.834-0.880) |
| Early Decliners | Gradual decline | 1,007 | 0.600 (0.562-0.637) | 0.790 (0.759-0.820) | 0.800 (0.769-0.829) |
| Early Decliners | Stable high | 1,058 | 0.765 (0.733-0.795) | 0.911 (0.892-0.929) | 0.922 (0.902-0.939) |
| Stable moderate | Gradual decline | 618 | 0.562 (0.518-0.607) | 0.593 (0.548-0.638) | 0.586 (0.544-0.628) |
| Stable moderate | Stable high | 669 | 0.647 (0.607-0.691) | 0.715 (0.676-0.754) | 0.727 (0.690-0.765) |
| Gradual decline | Stable high | 631 | 0.674 (0.631-0.715) | 0.717 (0.676-0.758) | 0.751 (0.713-0.788) |

Values are conditional pairwise AUROC (95% CI). For each row, evaluation was restricted to patients in the two indicated trajectory classes. The existing four-class probabilities were renormalized; no separate binary models were fitted. CPAP predictors summarize calendar days 1-7 beginning with the first night of use. AUROC indicates area under the receiver-operating-characteristic curve; CI, confidence interval.

### Supplement S17. Phase 4 held-out AUROC for Phase 2 outcomes across full and reduced-feature model specifications.

| Model specification | Month 12: mean use ≥4 hours/night | Month 12: mean use ≥2 hours/night | Month 12: any use | CMS 90-day adherence |
|---|---|---|---|---|
| Full baseline | 0.678 (0.657–0.697) | 0.676 (0.656–0.694) | 0.658 (0.639–0.678) | 0.653 (0.633–0.673) |
| Full baseline + CPAP | 0.810 (0.793–0.826) | 0.816 (0.800–0.831) | 0.799 (0.783–0.815) | 0.899 (0.889–0.910) |
| CPAP-data only | 0.787 (0.770–0.804) | 0.791 (0.774–0.807) | 0.778 (0.761–0.795) | 0.893 (0.882–0.904) |
| Parsimonious baseline | 0.644 (0.624–0.664) | 0.641 (0.621–0.660) | 0.627 (0.607–0.646) | 0.651 (0.631–0.671) |
| Parsimonious + CPAP | 0.804 (0.787–0.820) | 0.809 (0.793–0.824) | 0.793 (0.777–0.809) | 0.899 (0.888–0.910) |
| Questionnaire-Sleep-Study baseline | 0.672 (0.651–0.691) | 0.659 (0.638–0.677) | 0.658 (0.639–0.677) | 0.644 (0.623–0.663) |
| Questionnaire-Sleep-Study + CPAP | 0.808 (0.791–0.824) | 0.815 (0.799–0.829) | 0.800 (0.784–0.816) | 0.898 (0.888–0.909) |

Values are AUROC (95% CI) from the KPSC random held-out test set (n=2,982), using 2,000 stratified bootstrap resamples. Baseline models use pretreatment features; combined models add CPAP-data from days 1-7 beginning with the first positive-use night. AUROC indicates area under the receiver-operating-characteristic curve; CI, confidence interval.

### Supplement S18. Phase 4 held-out one-versus-rest AUROC for Phase 3 trajectory prediction across full and reduced-feature model specifications.

| Model specification | Macro average | Early Decliners | Stable moderate use | Gradual decline | Stable high use |
|---|---|---|---|---|---|
| Full baseline | 0.619 (0.605–0.632) | 0.667 (0.647–0.685) | 0.584 (0.559–0.610) | 0.533 (0.506–0.561) | 0.691 (0.667–0.713) |
| Full baseline + CPAP | 0.770 (0.759–0.781) | 0.871 (0.859–0.884) | 0.691 (0.668–0.714) | 0.657 (0.633–0.681) | 0.859 (0.843–0.874) |
| CPAP-data only | 0.756 (0.744–0.767) | 0.861 (0.848–0.873) | 0.673 (0.649–0.696) | 0.646 (0.621–0.669) | 0.843 (0.826–0.860) |
| Parsimonious baseline | 0.600 (0.587–0.613) | 0.642 (0.624–0.661) | 0.559 (0.532–0.585) | 0.542 (0.514–0.569) | 0.657 (0.634–0.680) |
| Parsimonious + CPAP | 0.767 (0.756–0.778) | 0.869 (0.856–0.881) | 0.683 (0.661–0.706) | 0.660 (0.636–0.683) | 0.856 (0.840–0.871) |
| Questionnaire-Sleep-Study baseline | 0.610 (0.597–0.624) | 0.660 (0.641–0.679) | 0.578 (0.552–0.604) | 0.522 (0.496–0.548) | 0.681 (0.657–0.703) |
| Questionnaire-Sleep-Study + CPAP | 0.768 (0.757–0.779) | 0.871 (0.859–0.883) | 0.689 (0.665–0.711) | 0.656 (0.632–0.679) | 0.856 (0.840–0.871) |

Values are one-versus-rest AUROC (95% CI) from the KPSC random held-out test set (n=2,981), using 2,000 stratified bootstrap resamples. Macro average is the unweighted mean across the four trajectory classes. Baseline models use pretreatment features; combined models add CPAP-data from days 1-7 beginning with the first positive-use night. AUROC indicates area under the receiver-operating-characteristic curve; CI, confidence interval.
